# The association between type of conception through medically assisted reproduction and childhood cognition: A Danish population-wide cohort study

**DOI:** 10.1101/2022.11.29.22282867

**Authors:** Peter Fallesen

## Abstract

Previous research has found that children conceived through medically assisted reproduction (MAR) on average has cognitive outcomes on par with or above naturally conceived children. However, previous work has been unable to consider the relationship at a full population level nor consider heterogeneity across type of MAR. We use all Danish live births in the years 2006-2009 (n=259,608) with indicator for MAR conceptions (n=13,554). The dependent variable is the within-year-and-grade standardized test scores carried out in second and third grade in primary schools. We compare the test scores for spontaneously conceived (SC) children and conceived through intrauterine insemination (IUI) and assisted reproductive technologies (ART). We estimate ordinary least squares regressions with a baseline model only adjusted for birthyear and models adjusted for birth-related confounders and socio-demographic family characteristics. At baseline, ART and IUI conceived children performed better in tests than SC peers. After adjusting, ART conceived performed worse than SC peer, and IUI conceived performed as well as SC peers and better than ART conceived. Results likely reflect differences in selection of potential parents into type of MAR as well as consequences of differences in fecundability.

---

With the ongoing rise in maternal age at birth across the global north^1^ and the suggested decline in human fecundity^2^, birth occurring following medically assisted reproduction (MAR) has consistently increased in the 21th century^3^. In 2018, 9.8 percent of births in the frontrunner country Denmark was children conceived following some type of MAR treatment^4^. With birth rates continuing to decline, MAR is poised to play an increasingly important role in upholding fertility levels^5^. Further, with rises in singlehood^6^ and same-sex families^7^ also occurring in recent decades, women whose sole barrier to motherhood is the absence of a male partner seek MAR treatment for different reasons than women trying but failing to conceive.

The growth in MAR birth has motivated an expanding literature that considers the consequences of MAR conceptions for children’s wellbeing and development. Children conceived through MAR have poorer birth outcomes^8,9^, but studies of later life wellbeing and cognitive ability generally find that association with cognitive ability is null or positive^10–17^. However, whereas studies using population wide data materials have been conducted for the association between MAR conception and birth outcomes^8,9^, the literature on cognitive ability and wellbeing has tended to be conducted on more selective cohort studies, which raises question of external validity of the findings^16^. The few population-wide studies conducted are somewhat dated and focus on more rare events, such as mental disorders^18,19^. No previous study has been able to examine the population wide association between MAR conception and cognitive ability, nor considered the role of type of mar treatment^16^.

The study has two aims. First, it presents evidence of relationship between MAR conception and mid-childhood cognitive development using population-wide data from four Danish birth cohorts born 2006-2009 (n=259,608). Denmark provides the optimal case study for considering population wide consequences of MAR births. From 2006, MAR treatment was made available in Denmark to all women, regardless of sexual orientation and partnership status. Further, public funding is available for three cycles of treatment insofar childlessness is due to infertility, it is the woman’s first children with present partner, body mass index (BMI) was below 30, the woman was younger than 40, and the woman was considered able to function as a parent. Thereby, financial barriers are lower in Denmark, also regardless of sexual orientation and partnership status. Thus, the well-established social gradient in likelihood of having children through MAR^20^ may be less steep in Denmark. Second, the study considers heterogeneity in cognitive across types of MAR, acknowledging that this heterogeneity might show important differences because model of conception^16^. We distinguish between conceptions following assisted reproductive technologies (ART), which includes all treatments in which either eggs or embryos are handled, and conceptions following intrauterine insemination (IUI). As a measure of cognitive development, we use results from low stakes standardized test administered in all Danish public schools and some private schools at second grade (test score available for all four cohorts, covers 77.5% of cohort members) and third grade (test score available for cohorts born 2006-2008, covers 76.9% of cohort members). Combining birth, population, educational, and income registers allow for a rich set of birth-related and socioeconomic controls.

## Method

### Sample description

We use four birth cohorts consisting of all live born Danish children born between January 1, 2006 to December 31, 2009 drawn from the Danish Medical Birth Register^21^, which includes all births in hospitals and from in-home deliveries. The data includes information on the unique personal identifiers of mother and child, as well as information on maternal and offspring health. Through the unique maternal identifier, we can link mothers to the Danish National Register of assisted reproductive technology^22^, which includes information on type and timing of all MAR treatments carried out in Denmark at public and private clinics. To obtain information on sociodemographic and -economic characteristics, we further link mothers to the Danish Population^23^, education^24^, and income^25^ registers. Finally, we link children to their second and third grade test scores using the register for the Danish National Tests^26^. The gross sample include 259,608 live births, of which 13,554 (5.2%) occurred within ten months of mother receiving any form of MAR treatment. We exclude children who died (n=1,165) or migrated (n=5,042) before age 9, which is the age children normally sit the grade 2 test. MAR conceived were 2.5% less likely to have a valid test score than naturally conceived (SC) children. However, after controlling for observable characteristics, the difference in the probability of being in the sample declined to 0.1% (see Table A1 in appendix).

Thus, ART conceived children are less likely to be in the estimation sample, but differences disappeared once adjusted for covariates.

### Variables

#### Outcome variable

The national tests were introduced in 2009 for all public schools (with an option to opt-in for private schools, although distinguishing between private and public schools are not possible in the data) and aimed to provide a uniform measure of child school performance in Denmark. The tests are conducted in the spring season and consist of a battery of different tests varying over grades. We focus on Danish reading comprehension test conducted in grade 2, and Math ability test conducted in grade 3. Tests are mandatory for all children enrolled in Danish public schools (private schools could opt-in), are adaptive, and are conducted on computers with an automatically generated result.^27^ The test results are not displayed in any formal school diplomas and have no direct implications for the child’s further school opportunities (there is no tracking or ability grouping in Danish primary or lower secondary schools). The test scores are standardized to mean 0 and standard deviation 1 within grade and year for all children sitting the test (and not just for the analytical sample).

#### Main independent variable of interest

Our indicator for conception following MAR treatment includes births that occurred within 10 months of a IUI and ART treatment cycle. ART includes intracytoplasmic sperm injection (ICSI), in vitro fertilization (IVF), vitrified-warmed blastocyst replacement (WBR)/frozen embryo replacement (FER), and oocyte donation (OD). IUI includes inseminations occurring fully inside the uterus. In case of multiple treatment occurred within 10 months of life birth, we use the last recorded treatment.

We adjust the analyses for a series of confounders known to correlate with probability of MAR treatment and children’s test scores. We divide these confounders into two groups: i) birth-related characteristics and ii) socioeconomic and -demographic characteristics. Birth related characteristics include birth year indicator, birthweight (categorical), multiple birth indicator (binary), parity (categorical, with multiple births coded with similar parity), maternal body mass index (BMI, categorical), maternal smoking during pregnancy (binary), and child gender (binary). These variables were obtained from the medical birth records. In case of missing information, a separate category was created capturing this. Socioeconomic and -demographic characteristics include whether maternal age (categorical), mother was married/cohabiting to/with a man/woman start of year of birth (binary), maternal level of education at start of year of birth (categorical), disposable income in €1k start of year of birth (continuous), maternal migration background (categorical), and whether child was young/old for grade when sitting the test (category). Grouping details and distribution of confounders are presented in detail the first part of the Results section.

### Statistical models

We estimate three linear regression models for each grade with standard errors clustered at the maternal level. First, a baseline model where we only adjust test scores for birth year. Second, a model where we adjust for birth year and birth-related confounder. Third, a model where we adjust for birth year, birth-related, and socioeconomic and -demographic confounders. We present separate estimates for ART and IUI, as well as estimates for the difference between ART and IUI. All analyses were conducted using Stata Statistical Software: Release 17 (StataCorp LP, College Station, TX).

## Results

### Descriptive results

Table 1 presents the descriptive statistics of the analytical sample by birth type for all births and by mode of conception (SC, ART, and IUI). There are fewer observations born in 2009 than previous years because we only have grade 3 test scores for birth cohorts 2006-2008. For both type of MAR, children are more likely to be born in the later of the included birth years than SC children. ART conceived children are less likely to be male compare to SC children, whereas IUI conceived children are more likely to be male. MAR children also have lower birthweight. 17.2% of ART and 12.8% of IUI conceived had birthweights below 2500 grams, whereas only 4.2% of SC conceived had birthweight below 2500 grams. Further. ART conceived were a factor of 10 (31%) more likely to be part of a multiple birth than SC children (3.1%), and IUI children (21.4%) a factor of 7.

**Table 1:** Descriptive statistics across conception mode

|  | ART | IUI | SC | Total |
| --- | --- | --- | --- | --- |
| Grade =2 | 58.9% | 60.1% | 57.1% | 57.2% |
| Child male=1 | 48.7% | 51.6% | 50.6% | 50.6% |
| Birth year |  |  |  |  |
| 2006 | 13.8% | 9.6% | 30.7% | 29.8% |
| 2007 | 34.7% | 32.4% | 28.8% | 29.0% |
| 2008 | 35.7% | 39.1% | 27.6% | 28.0% |
| 2009 | 15.8% | 18.9% | 12.9% | 13.1% |
| Birthweight |  |  |  |  |
| <1499 grams | 2.4% | 1.6% | 0.5% | 0.6% |
| 1500-2499 grams | 14.8% | 11.2% | 3.7% | 4.3% |
| 2500-3499 grams | 49.9% | 49.39% | 42.3% | 42.6% |
| 3500-4499 grams | 30.5% | 35.3% | 48.7% | 48.7% |
| 4500+ grams | 1.6% | 1.95% | 3.3% | 3.3% |
| Unknown | 0.9% | 0.6% | 1.6% | 1.6% |
| Multiple births | 31.0% | 21.4% | 3.1% | 4.3% |
| Maternal age |  |  |  |  |
| <18 years | NA | NA | 0.3% | 0.3% |
| 18-24 years | 1.6% | 2.3% | 12.0% | 11.5% |
| 25-29 years | 19.2% | 23.3% | 32.7% | 32.1% |
| 30-34 years | 45.3% | 44.1% | 37.4% | 37.8% |
| 35-39 years | 27.2% | 25.2% | 15.0% | 15.6% |
| 40+ years | 6.7% | 5.0% | 2.7% | 2.8% |
| Parity |  |  |  |  |
| 1 | 62.6% | 62.3% | 42.5% | 43.5% |
| 2 | 28.8% | 30.1% | 36.1% | 35.7% |
| 3 | 4.2% | 4.7% | 13.9% | 13.4% |
| 4+ | 1.5% | 0.7% | 4.7% | 2.9% |
| Unknown | 2.9% | 2.2% | 2.9% | 2.9% |
| Maternal BMI |  |  |  |  |
| <18.5 | 7.4% | 7.5% | 7.9% | 7.9% |
| 18.5-24.9 | 60.4% | 58.2% | 57.6% | 57.7% |
| 25.0-29.9 | 21.3% | 19.7% | 19.5% | 19.6% |
| 30.0-34.9 | 7.0% | 8.9% | 7.3% | 7.4% |
| 35.0-39.9 | 1.5% | 3.0% | 2.6% | 7.4% |
| 40+ | 0.5% | 0.9% | 1.3% | 1.3% |
| Unknown | 2.6% | 1.8% | 3.7% | 3.7% |
| Maternal smoking | 7.2% | 7.4% | 14.8% | 14.4% |
| Maternal partner =1 | 96.7% | 91.6% | 88.7% | 89.0% |
| Maternal education |  |  |  |  |
| Lower secondary | 9.7% | 9.6% | 22.1% | 21.5% |
| Upper secondary | 38.4% | 37.0% | 40.3% | 40.2% |
| Tertiary | 51.9% | 53.4% | 37.6% | 38.3% |
| Maternal origin |  |  |  |  |
| Danish native | 92.4% | 93.2% | 87.5% | 87.8% |
| 1st gen immigrant | 6.9% | 5.8% | 10.8% | 10.6% |
| 2nd gen immigrant | 0.5% | 0.9% | 1.3% | 1.2% |
| Unknown | 0.1% | 0.1% | 0.4% | 0.4% |
| Disposable income | 24.946 (10.478) | 24.947 (9.612) | 21.480 (31.115) | 21.650 (30.439) |
| Child's age for grade |  |  |  |  |
| Young for grade | 1.1% | 1.6% | 1.7% | 1.7% |
| Aged for grade | 91.2% | 90.7% | 91.7% | 91.6% |
| Old for grade | 7.7% | 7.7% | 6.6% | 6.7% |
| Observations | 11,236 | 5,947 | 334,710 | 351,893 |

SC: Spontaneously conceived. ART: Assisted reproductive technology. IUI: Intrauterine insemination. BMI: Body mass index. Lower secondary education: ISCED < 3. Upper secondary education: ISCED = {3,4}. Tertiary education: ISCED > 4. ISCED: International Standard Classification of Education. Income measured in €1,000. Standard error in parentheses.

In term of birth-related maternal characteristics, ART mothers were older (33.9% aged 35 or older) than IUI mothers (30.2% aged 35 or older), who in turn were older that mother of SC children (17.7% aged 35 or older). Both types of MAR conceived children were also more likely to be their mother’s first birth event (ART: 62.6%, IUI 62.3%, SC 42.5%). Mothers of ART children had lower BMI (89.1% had BMI < 30) than mothers of SC children (85.0% had BMI < 30), whereas differences between SC and IUI (85.4% had BMI < 30) were more muted. MAR mothers were half as likely to smoke during pregnancy (ART: 7.2%, IUI 7.4%, SC 14.8%). In terms of socioeconomic and -demographic characteristics, MAR mothers had higher levels education (51.9% of ART had a tertiary degree, 53.4% of IUI, and 37.6% for SC), where less likely to have a migration background (92.4% of ART was Danish native, 93.2% of IUI, and 87.5% for SC), and had higher disposable income (post-tax and -transfers). Last, MAR conceived children was slightly more likely to start school later, and thus being old-for-grade (ART: 7.7%, IUI 7.7%, SC 6.6%).

### Regression results

Table 2 shows the coefficients [and 95% confidence intervals (CI)] from regressing the within-grade-and-year population standardized test score on mode of conception with SC as the reference category per grade. The full model results are presented in Supplementary Table A2-A3, and Figure 1 presents the predictive margins predicted at the mean value of adjustment variables across conception modes, models, and grade of test. It should be noted that the test scores are standardized within the full population of all children taking the test, and not just children born in Denmark within the birth years we consider, which is why the predictive margins in Figure 1 tend to all be larger than 0.

**Table 2:**
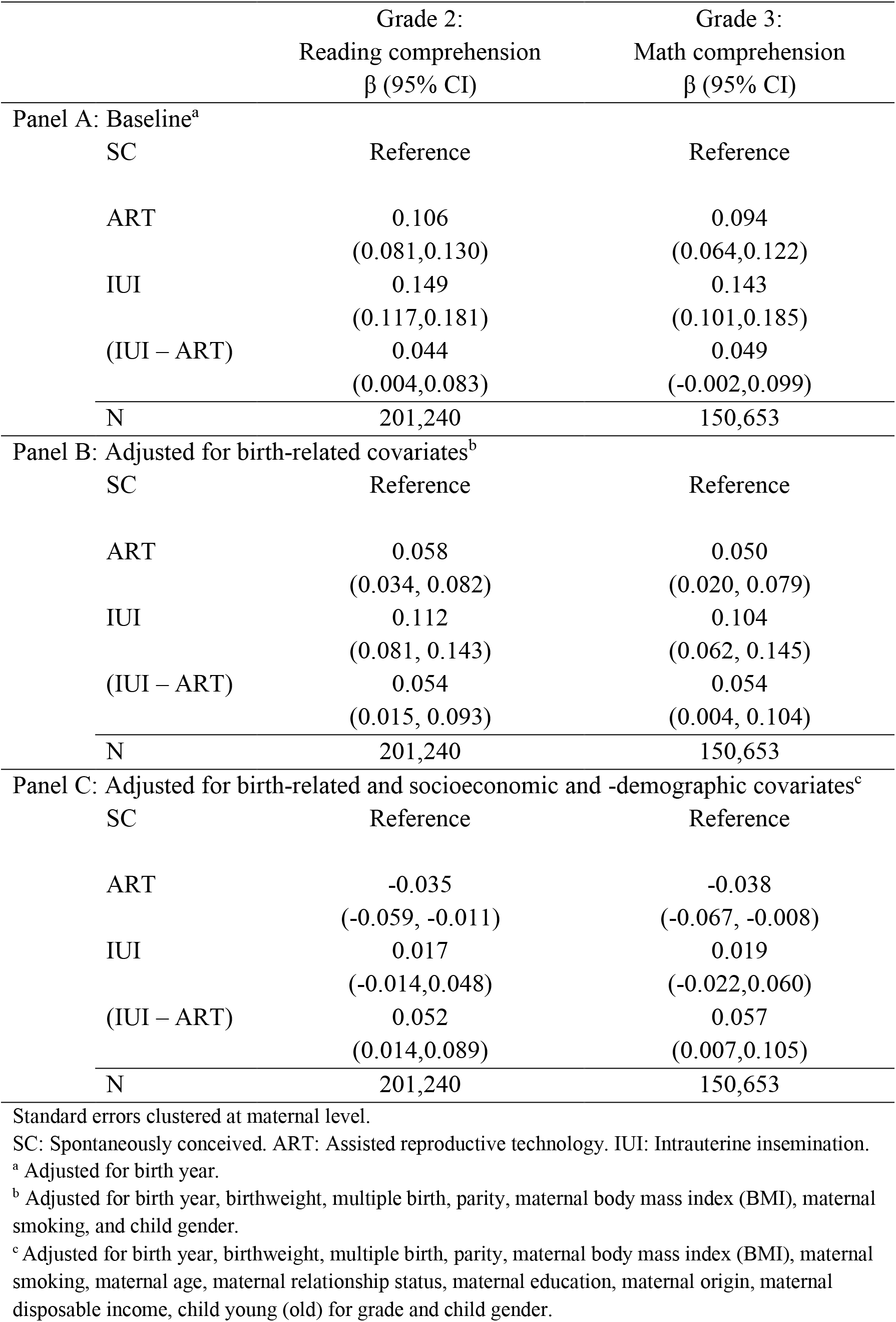
Linear models regressing test scores (standardized) on mode of conception

**Figure 1:**
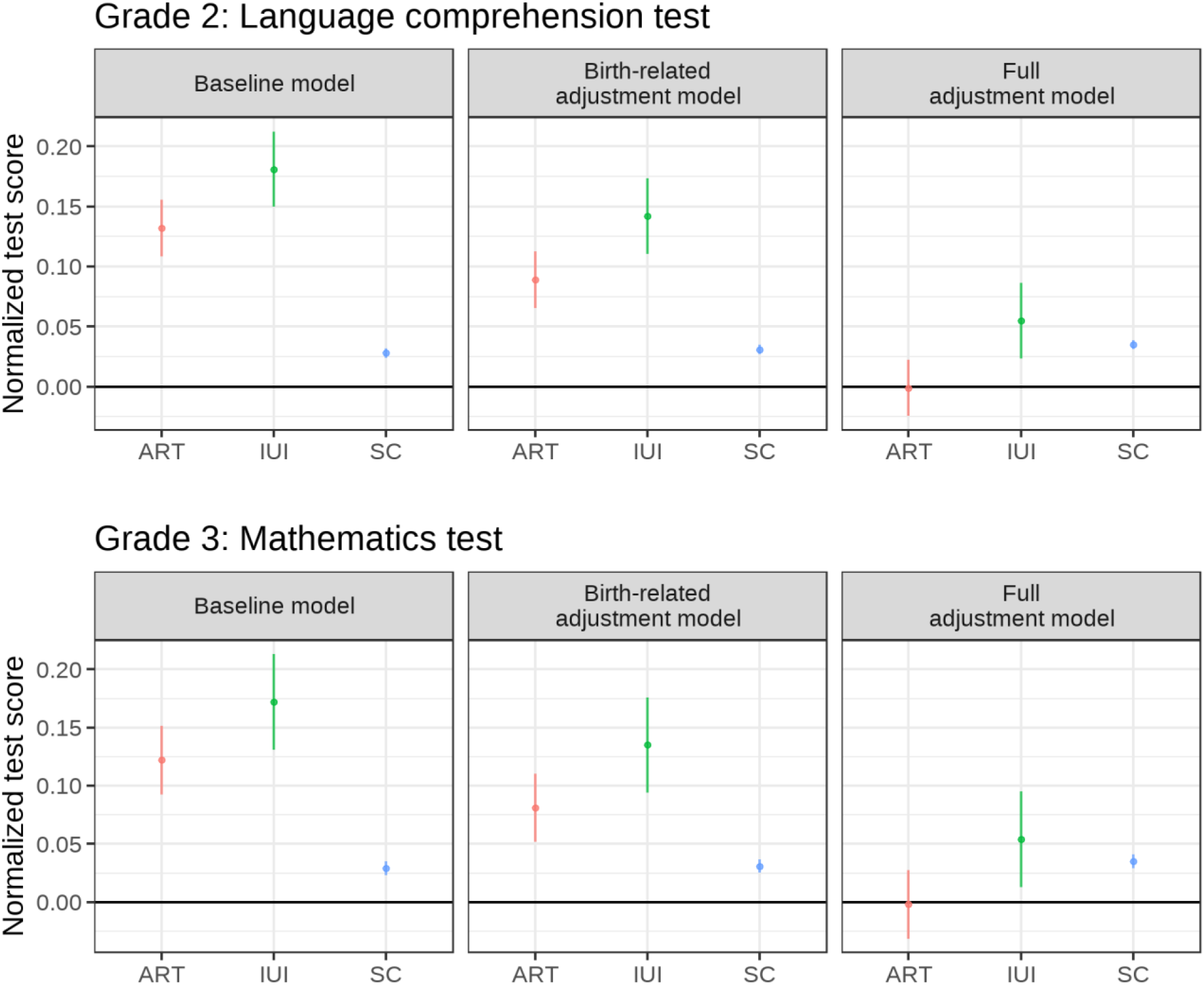
Predictive margins for normalized test scores and 95% confidence intervals by mode of conception Standard errors clustered at maternal level. SC: Spontaneously conceived. ART: Assisted reproductive technology. IUI: Intrauterine insemination. Baseline: Adjusted for birth year. Birth-related adjustment: Adjusted for birth year, birthweight, multiple birth, parity, maternal body mass index (BMI), and maternal smoking, child gender. Full adjustment: Adjusted for birth year, birthweight, multiple birth, maternal age, parity, maternal body mass index (BMI), maternal smoking, maternal relationship status, maternal education, maternal origin, maternal disposable income, child young (old) for grade and child gender.

Panel A reports the estimates from the baseline model only adjusted for birth year (left panels in Figure 1). ART and IUI conceived children have higher standardized test scores the SC children at both grade 2 (*β*^*ART*^=0.106, 95% CI: 0.081, 0.130; *β*^*IUI*^=0.149, 95% CI: 0.117, 0.181) and grade 3 (*β*^*ART*^=0.094, 95% CI: 0.064, 0.122; *β*^*IUI*^=0.143, 95% CI: 0.101, 0.185). Further, IUI conceived children also have higher standardized test scores than ART conceived children (*β*^*IUI*−*ART*^=0.044, 95% CI: 0.004, 0.083; *β*^*IUI*−*ART*^=0.049, 95% CI: -0.002, 0.099).

After adjusting for birth-related characteristics in Panel B (middle panels in Figure 1), the higher standardized test scores are somewhat attenuated but both ART and IUI conceived children still have higher standardized test scores than SC children [grade 2 (*β*^*ART*^=0.058, 95% CI: 0.034, 0.082; *β*^*IUI*^=0.112, 95% CI: 0.081, 0.143); grade 3 (*β*^*ART*^=0.050, 95% CI: 0.020, 0.079; *β*^*IUI*^=0.104, 95% CI: 0.062, 0.145)], and the difference between ART and IUI children slightly increase (grade 2: *β*^*IUI*−*ART*^=0.054, 95% CI: 0.015, 0.093; grade 3: *β*^*IUI*−*ART*^=0.054, 95% CI: 0.004, 0.104).

Last, after adjusting also for socioeconomic and -demographic characteristics in Panel C (right panels in Figure 1), ART conceived children have lower test scores than SC children in both grade 2 (*β*^*ART*^=-0.035, 95% CI: -0.059, -0.011) and grade 3 (*β*^*ART*^=-0.038, 95% CI: -0.067, -0.008). For IUI conceived children, there is no detectable difference between them and NC children at grade 2 (*β*^*IUI*^=0.017, 95% CI: -0.014, 0.048) and grade 3 (*β*^*IUI*^=0.019, 95% CI: -0.022, 0.060). The difference between ART and IUI conceived children remain similar to those found in previous models (grade 2: *β*^*IUI*−*ART*^ =0.052, 95% CI: 0.014, 0.089; grade 3: *β*^*IUI*−*ART*^=0.057, 95% CI: 0.007, 0.105).

As a first sensitivity analysis, we divide the sample by whether mother had a university degree or not to take the educational gradient in MAR usage into account. Figure 2 present the predicted margins from the fully adjusted model (full set of parameters shown in appendix Table A4). Differences in conception are larger among mothers with education below university than among those with any university education. However, differences between modes of conception are vastly overshadowed by differences by maternal education. Second, we limited the sample to only include singleton births, because multiple birth are more common following MAC conception and found to have lower academic achievement^28^. Table A5-A6 in supplementary materials reports the regression coefficients from this sensitivity analysis. The patters across conception types remained the same, although the difference between ART and IUI conceived children attenuated. As a third sensitivity analysis, we limited the sample to only include first births. This was done because first births are overrepresented among MAR conceived (see Table 1) and birth order is associated with test score advantage^29^. Table A7-A8 in supplementary materials reports the results from this sensitivity analysis. The patters across conception types remained the same, and the difference between ART and IUI conceived children were even larger.

**Figure 2:**
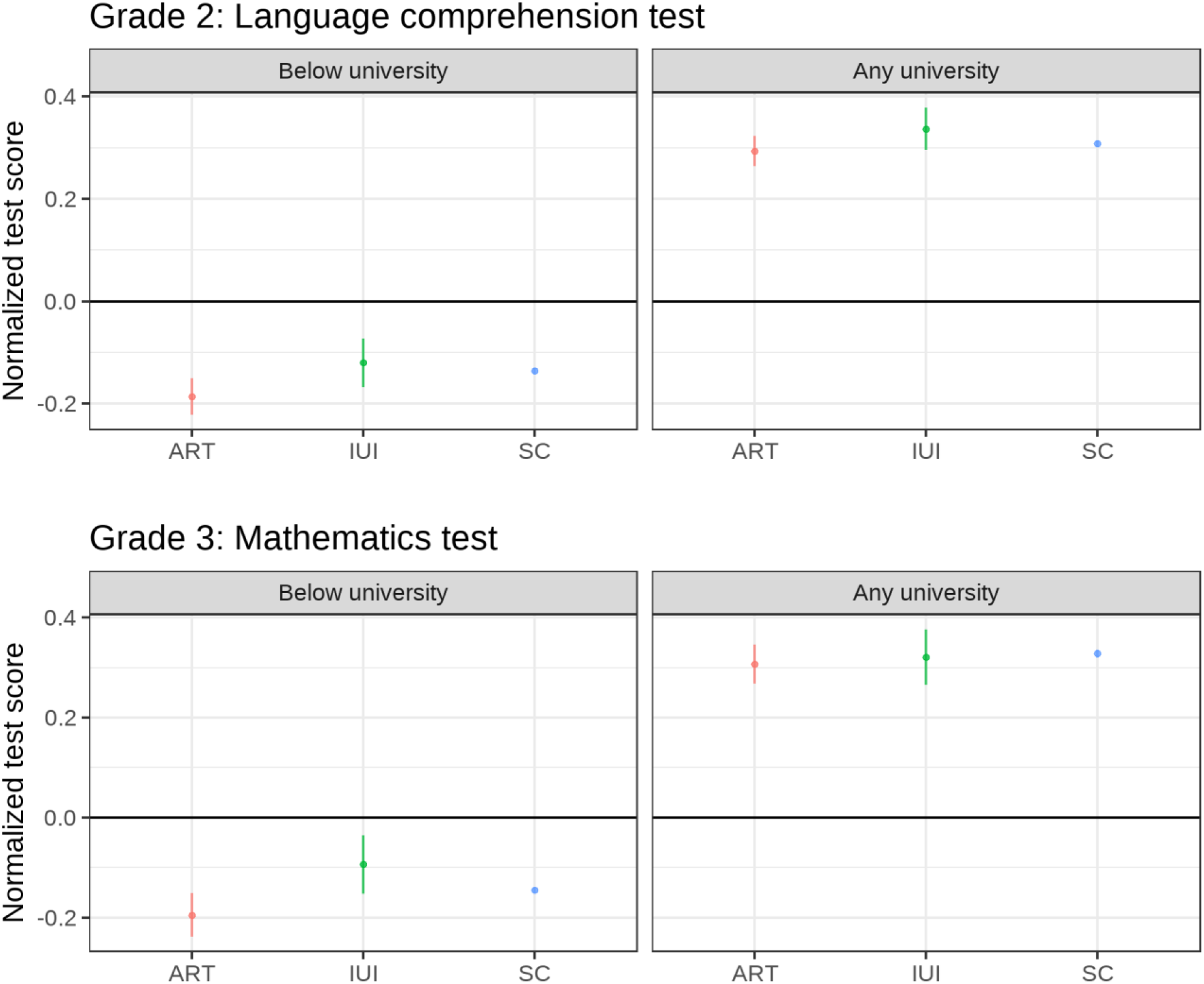
Predictive margins for normalized test scores and 95% confidence intervals by mode of conception Standard errors clustered at maternal level. SC: Spontaneously conceived. ART: Assisted reproductive technology. IUI: Intrauterine insemination. Full adjustment: Adjusted for birth year, birthweight, multiple birth, maternal age, parity, maternal body mass index (BMI), maternal smoking, maternal relationship status, maternal education, maternal origin, maternal disposable income, child young (old) for grade and child gender.

## Discussion

In this study, we used population wide data for four Danish birth cohorts to study whether the association between conception type (ART, IUI, or SC) and children’s test scores in grade 2 and 3. Our results showed that although ART and IUI conceived children at baseline had higher test scores than SC children, these differences disappeared once adjusting for birth, socioeconomic, and sociodemographic characteristics, with ART conceived children found to perform below both SC and IUI children.

Our study extends the literature on MAR conceived children’s cognitive development by using population wide data from a context with high usage and easy availability of MAR treatment^30^ and smaller variation in early life care and school quality than most other countries^31,32^, with previous work predominantly using data from settings with lower MAR uptake and more educational inequality, such as the UK^10,11^. By considering Denmark, where MAR conceived births now make up more than 8% of all birth, we have provided a vanguard view on the relationship between MAR conception and children’s academic achievement.

The second key finding is that ART conceived children consistently had lower test scores than IUI conceived, even in fully adjusted models. A possible explanation, although not possible to examine in the present study, is that parents conceiving through ART have lower fecundability than those conceiving through IUI. This would also account for the lower sex ratio in ART conceived than IUI conceived children, which is known to decline under harsher conditions^33^, as well as the higher risk of low birthweight. It is further supported by ART mothers more often being in a relationship and older than IUI mothers, which suggests a longer period until successful conception. Recent work by *Magnus et al*.^34^ has linked parental subfecundability to slower neurodevelopmental difficulties and delays in children, which may explain the gradient within MAR treatment in children’s test scores.

The present study has several limitations worth noting. First, we only observe MAR conceptions that occurred in Denmark. Thus, women who have run out of cycles or for other reasons were note eligible for public funding for MAR may have conceived at clinics outside Denmark. These births would be misclassified as SC. However, the bias this would cause would in any estimated differences between MAR and non-MAR conceptions would likely trend towards 0. Second, a key variable of interest we do not observe is time from beginning to try to conceive to successful conception, which would allow for a more thorough consideration of the role of fecundability. Third, MAR conceived children were a little less likely to not participate in the test. Last, we do not observe children’s cognitive ability earlier in childhood, leaving more time for environmental influences to make up for any initial developmental difficulties.

Nonetheless the study also has considerable strengths. First, it uses a nation-wide multi-cohort dataset, which is likely much less selected than other samples previously used in the literature studying cognitive ability, which in turn allowed us to also distinguish between type of MAR treatment. Second, we study a general outcome captured for all children (test scores) in a uniform a dispassionate way, rather than more rare neurodevelopmental outcomes, such as psychiatric diagnoses, which would be subject to clinicians’ discretion and might be influenced by knowledge about the way the child was conceived.

In conclusion, this study has contributed important new knowledge demonstrating that in the country with the highest MAR conception rates in the world, there exist differences in children’s educational achievement depending on mode of MAR conception. While not insubstantial, differences across type of MAR conception are vastly overshadowed by socioeconomic differences in educational achievement. Increase usage of ART may lead to more children with slightly lower test scores, but these differences are minor compared to differences along SES gradients.

## Data Availability

The data underlying this article cannot be shared publicly due to privacy concerns restricting availability of register data for research. The author can make aggregated data available, conditional on ethical vetting. The author accessed the individual-level data through Statistics Denmark online access system. If a researcher at a university or other research institution outside Denmark wishes to use these data, this may be accomplished by visiting a Danish research institution or by cooperating with researchers or research assistants working in Denmark. All scripts to generate data and results underlying the study are available as Supplementary Materials.

---

**Table A1:**
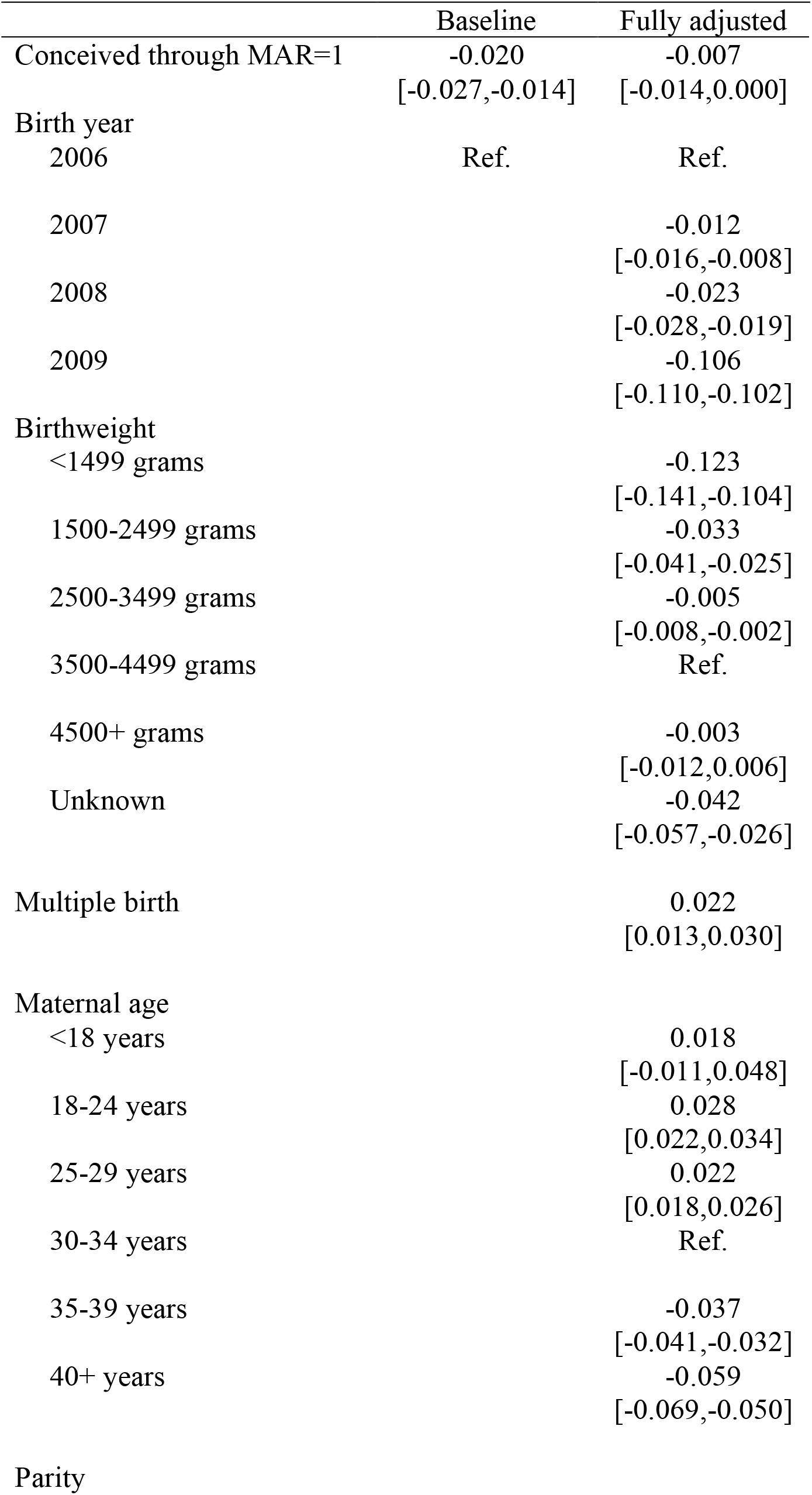
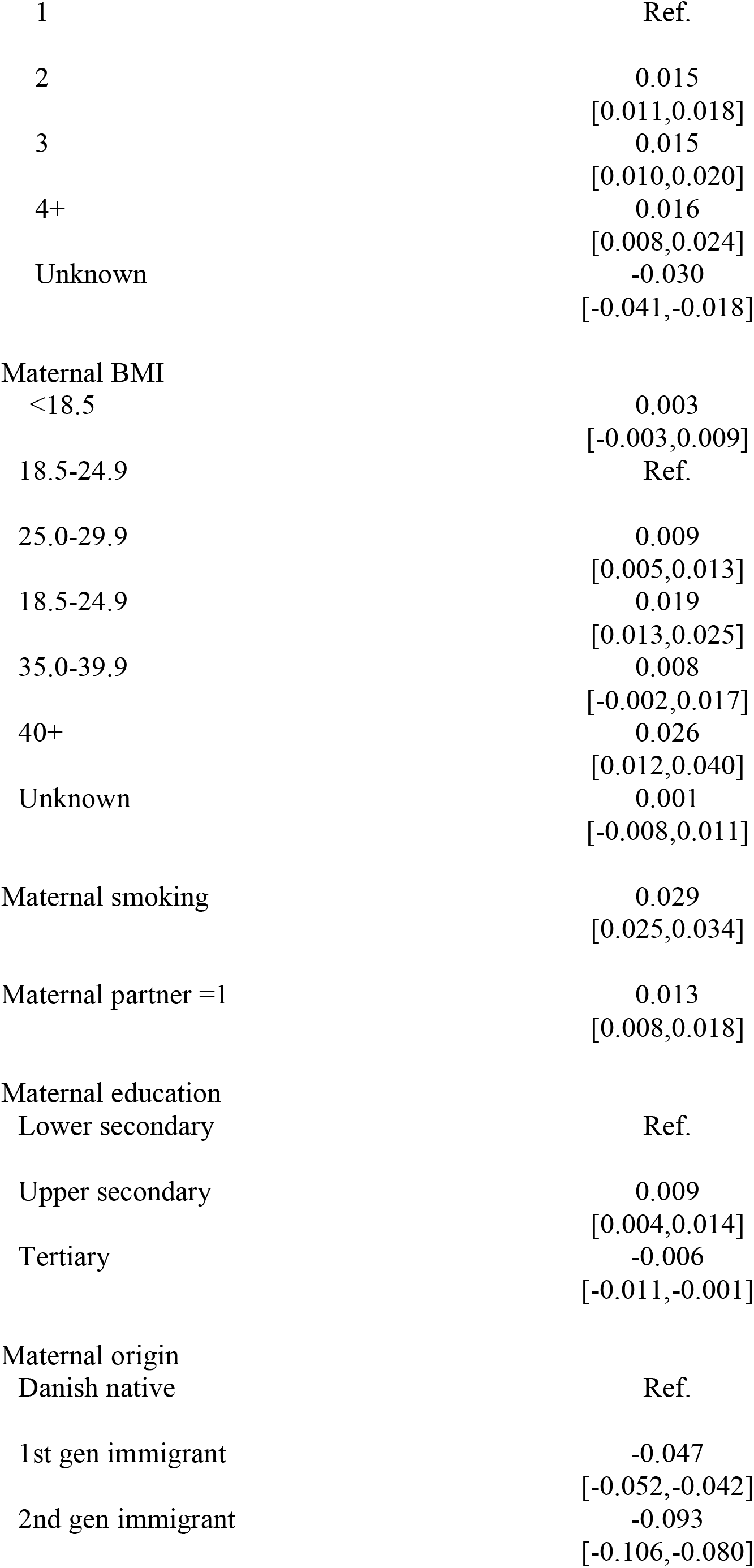
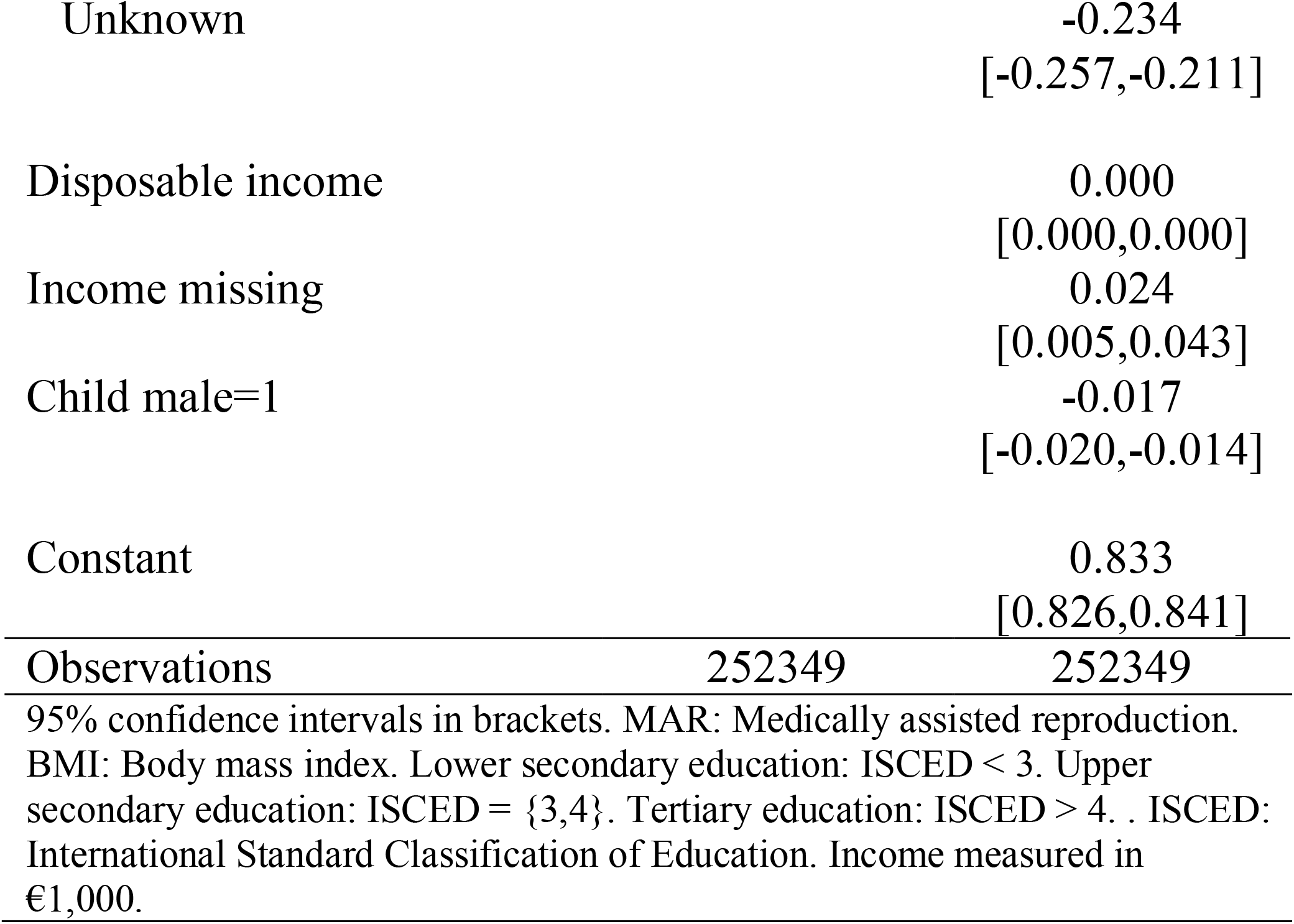
Linear model regressing being in the analytical sample on mode of conception and covariates

**Table A2:**
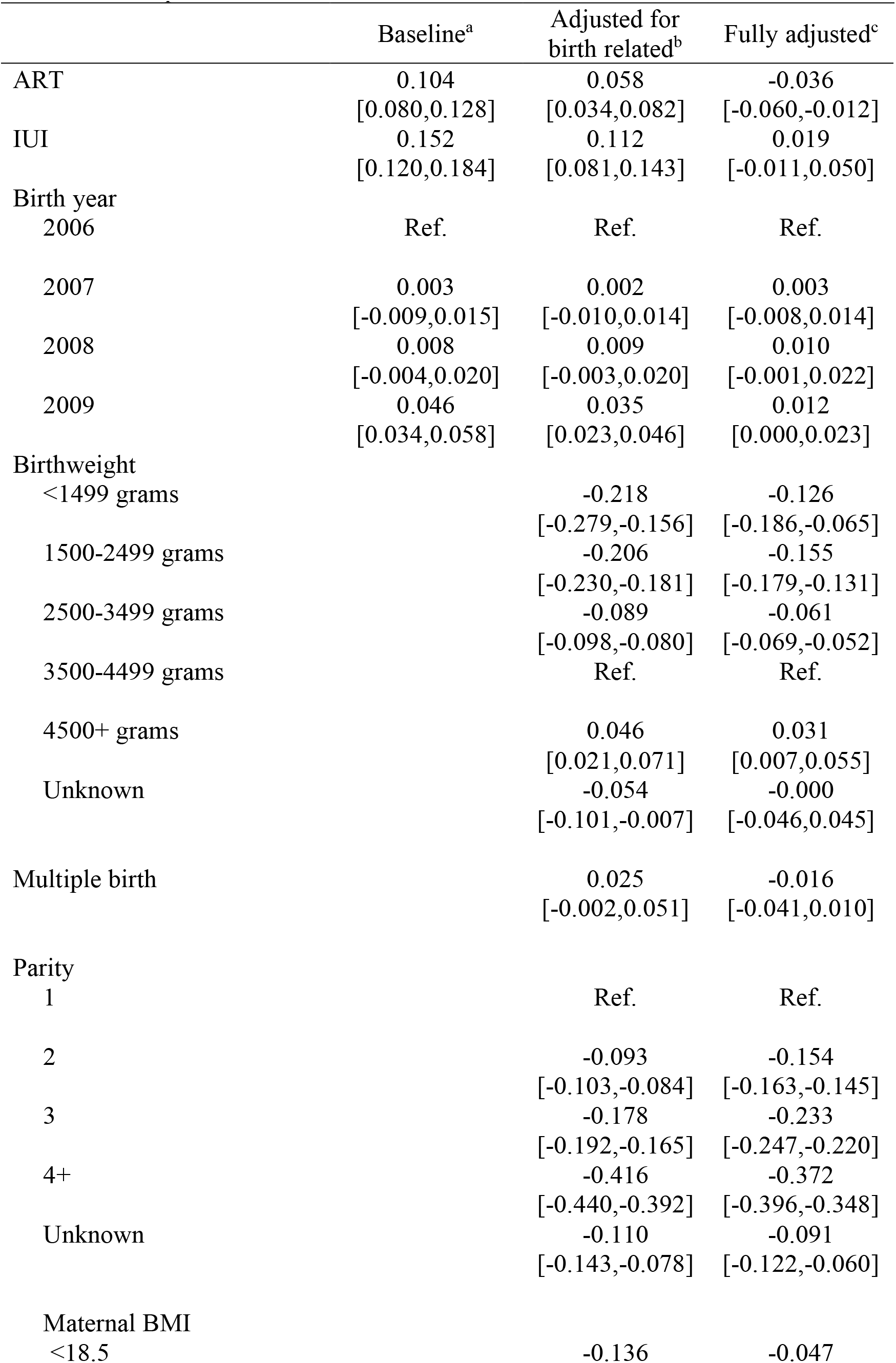
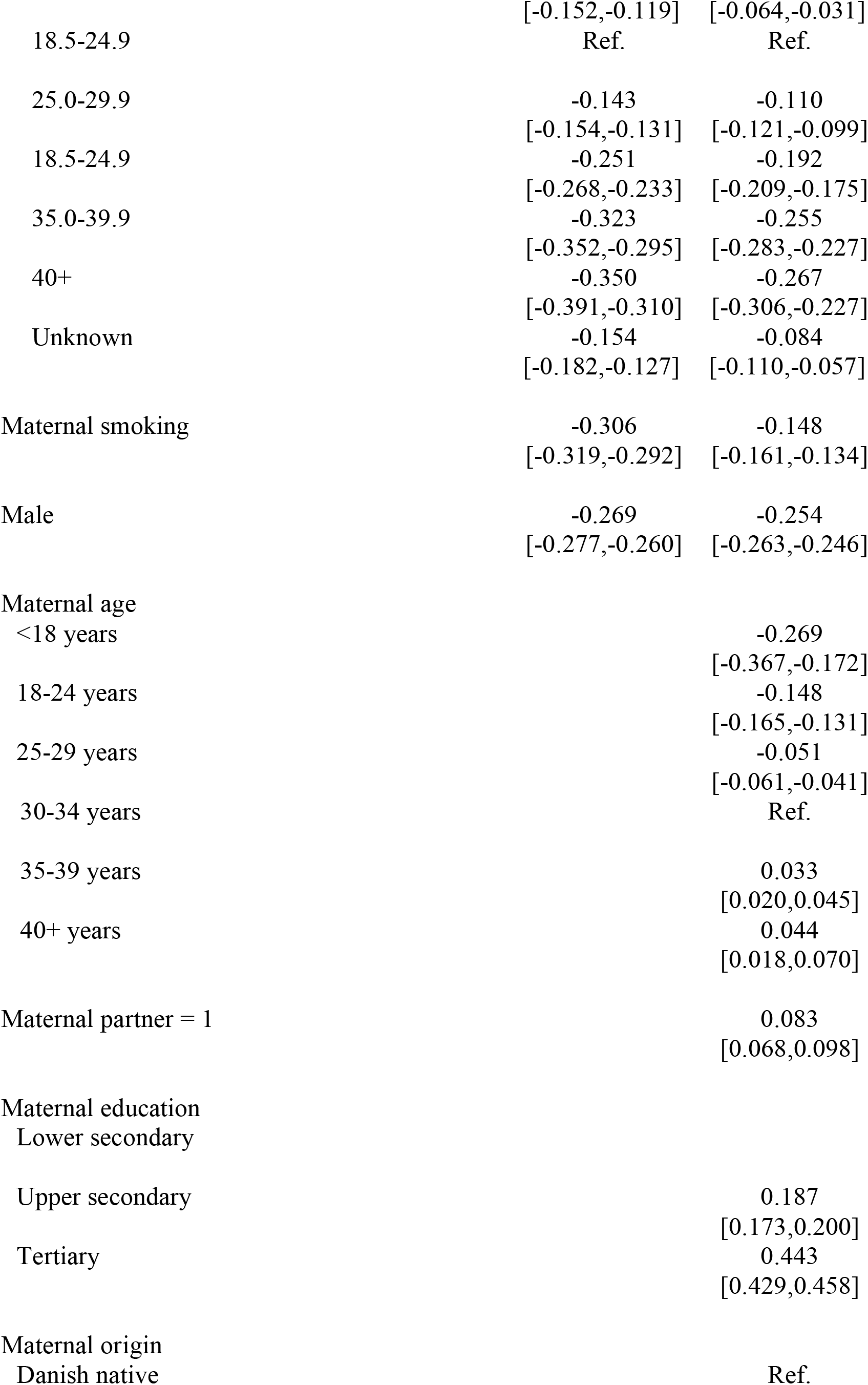
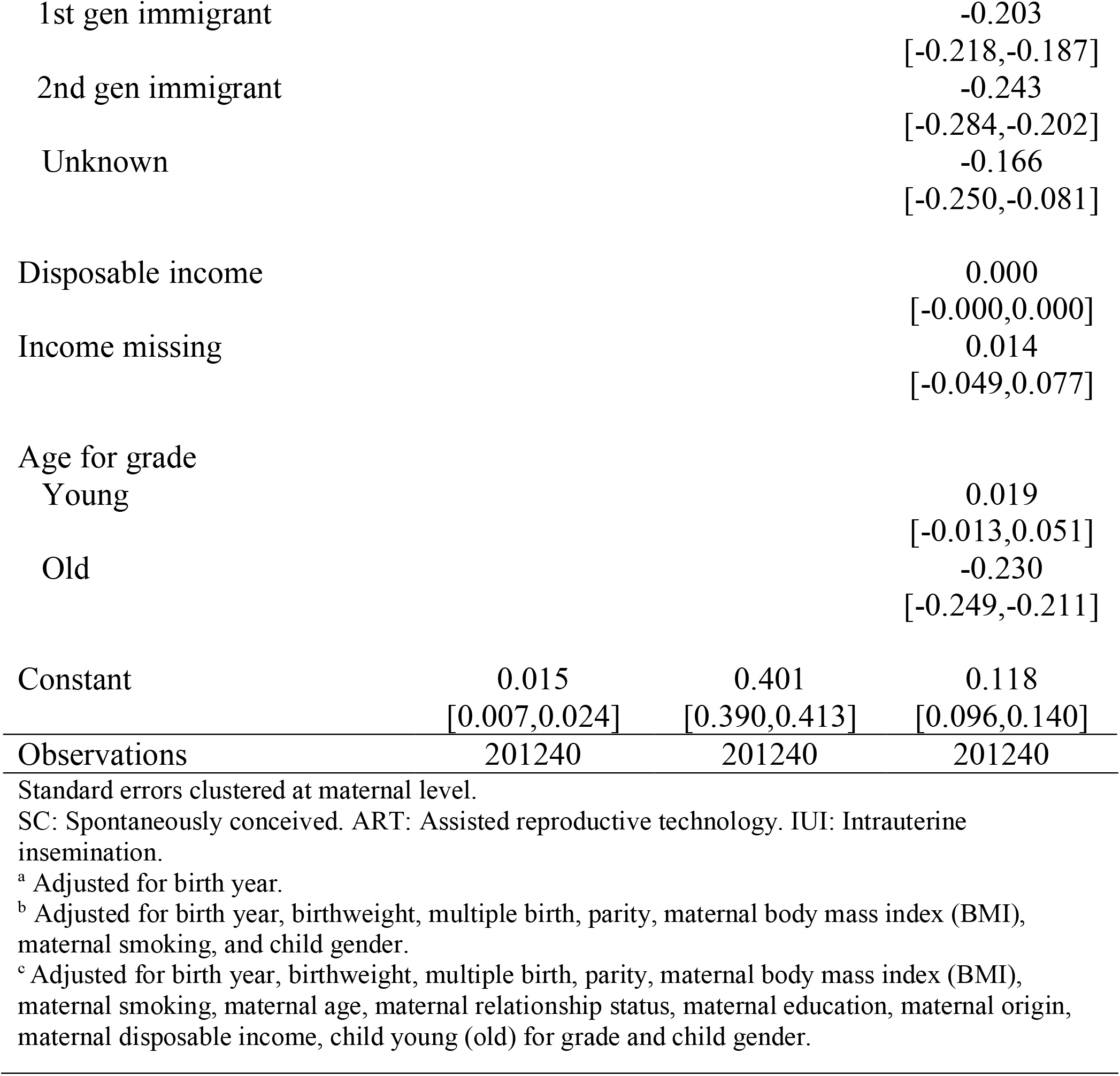
Linear model regressing second grade language comprehension score on mode of conception and covariates

**Table A3:**
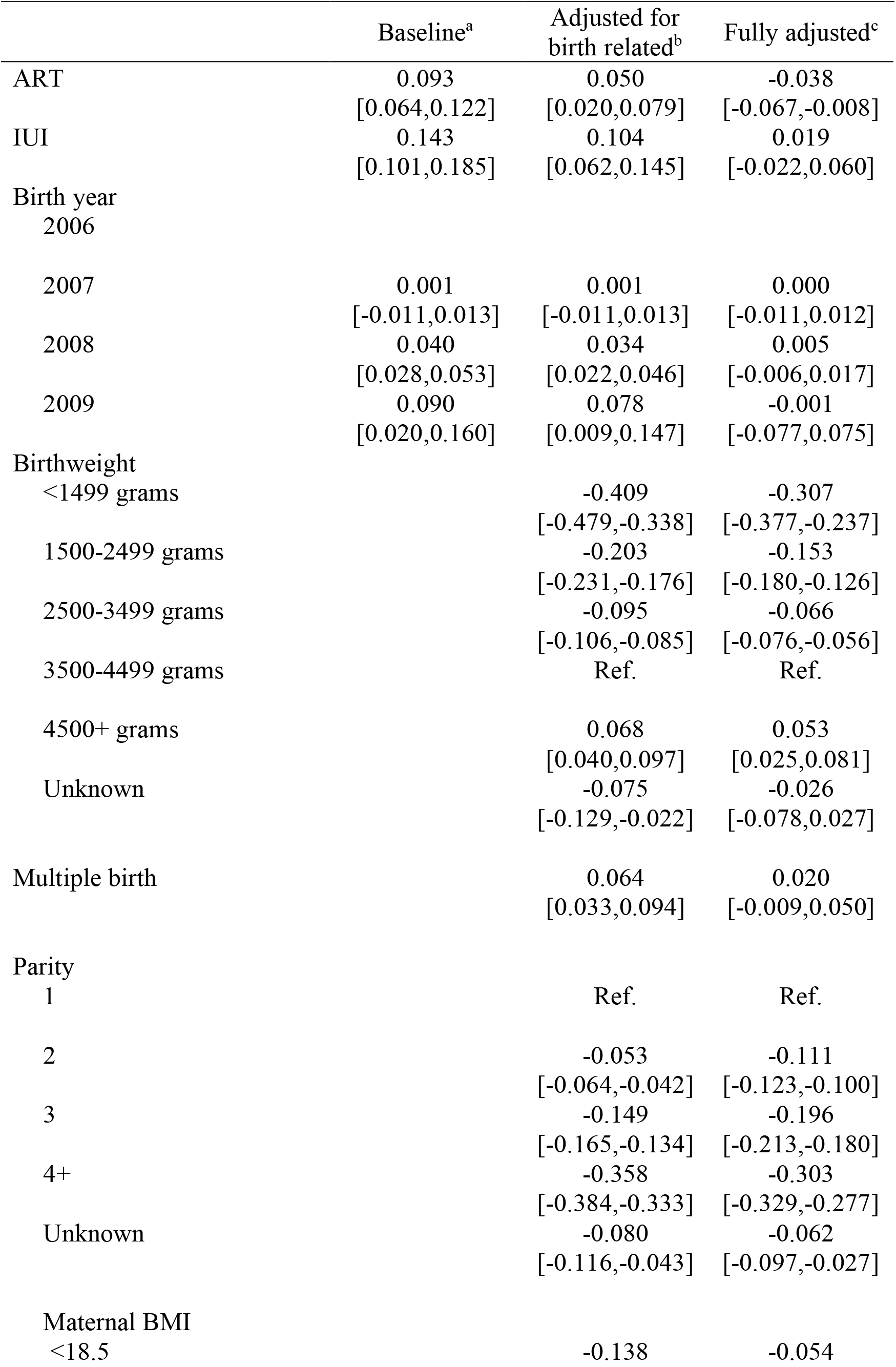
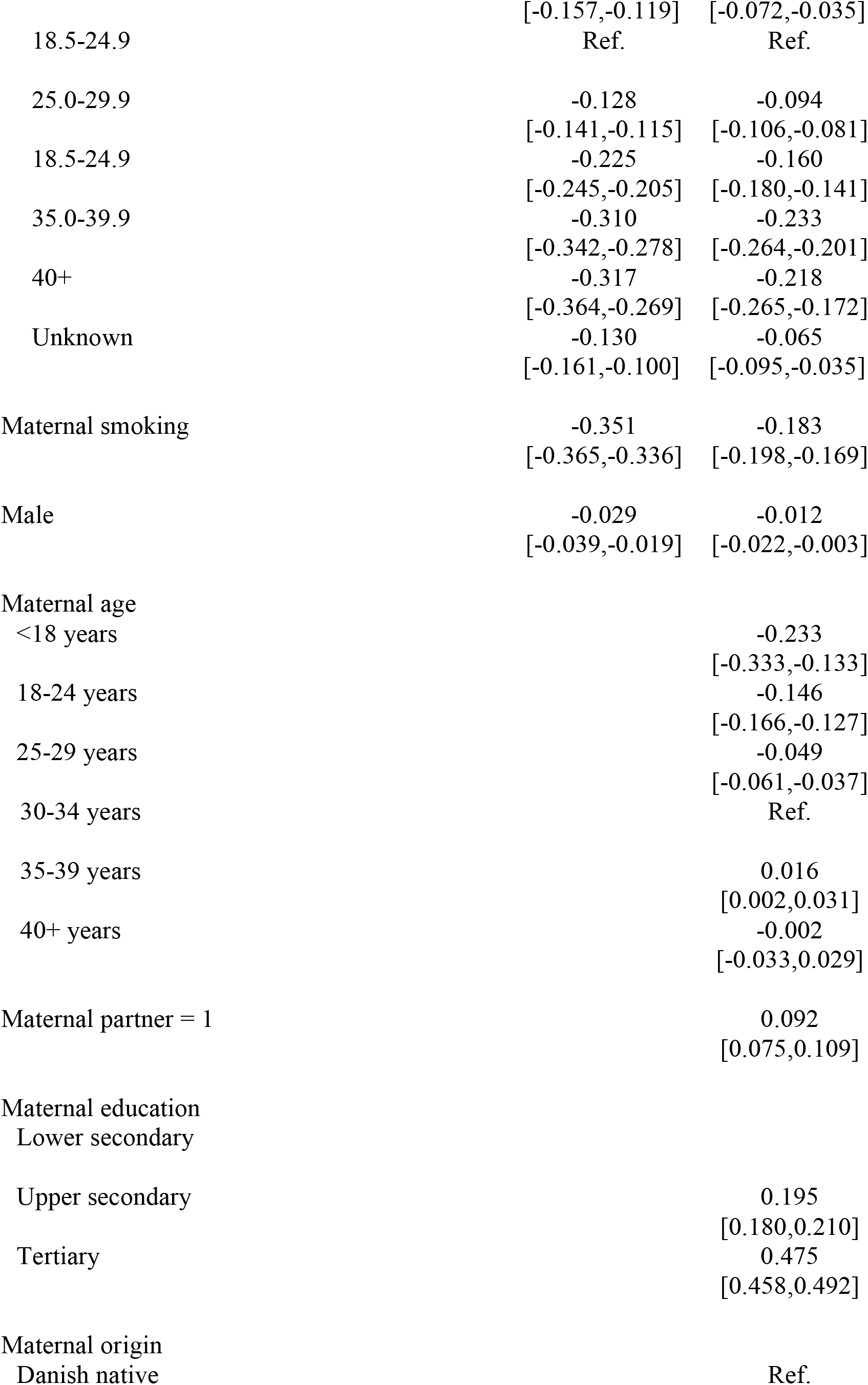
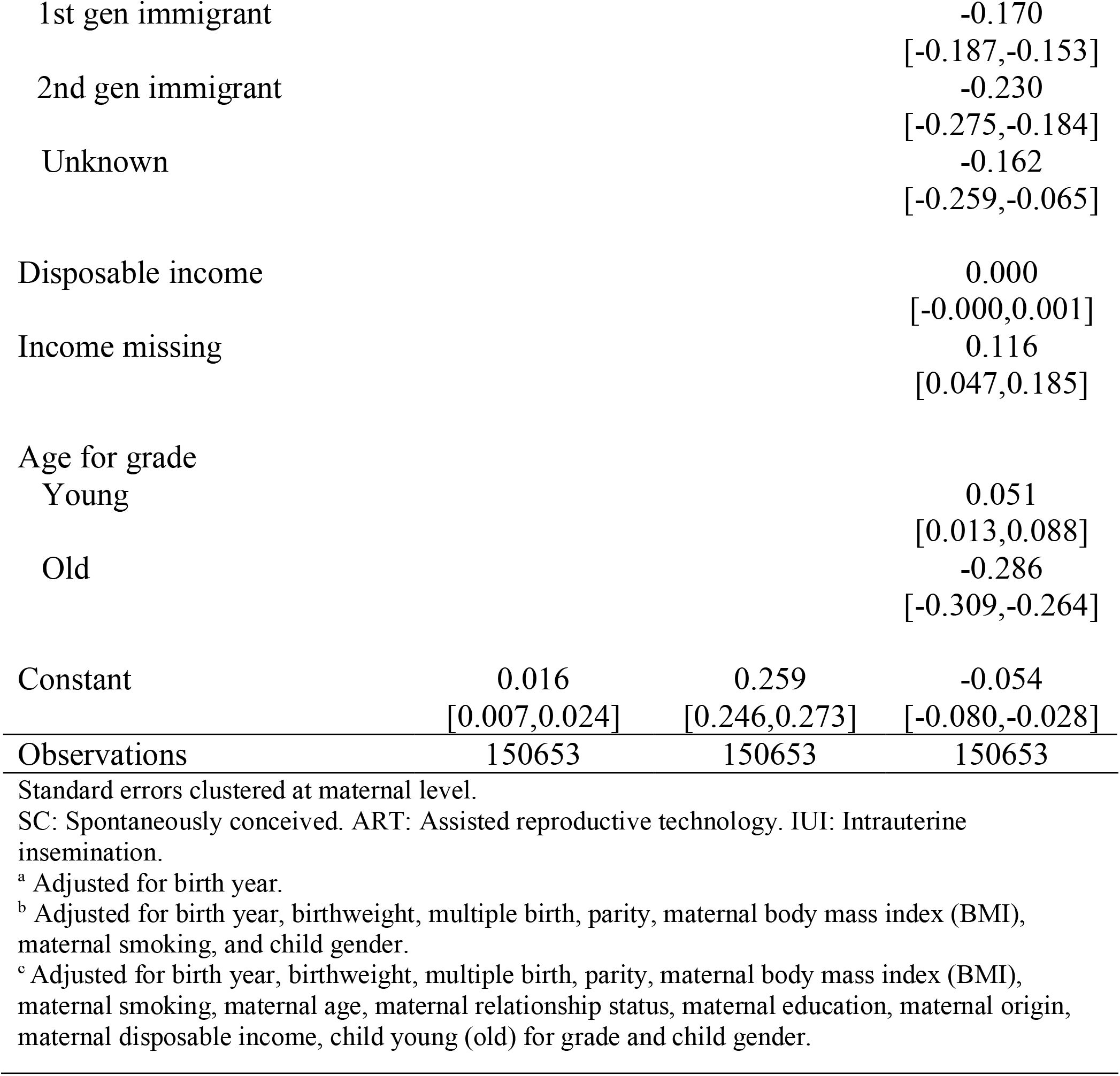
Linear model regressing third grade math score on mode of conception and covariates

**Table A4:**
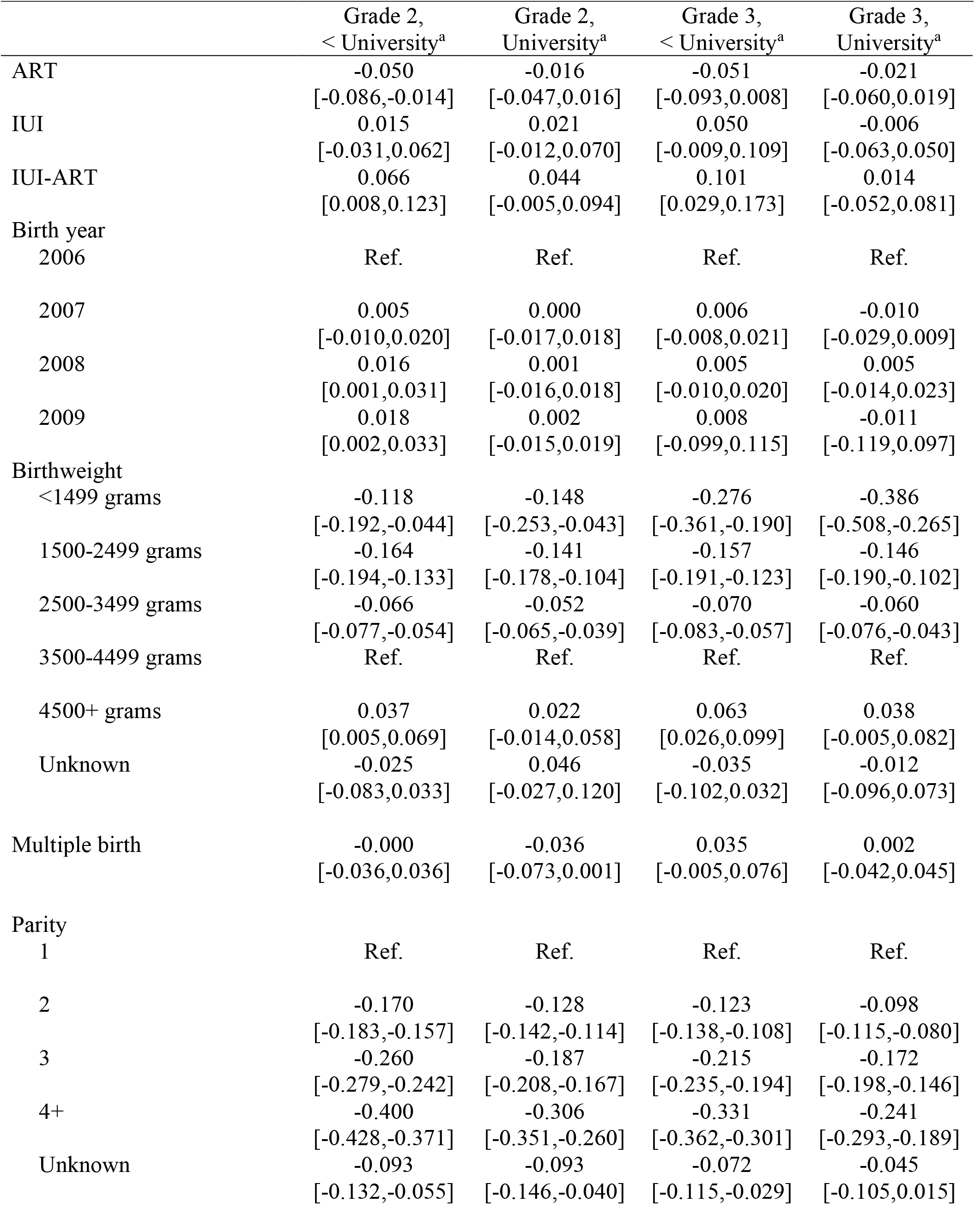
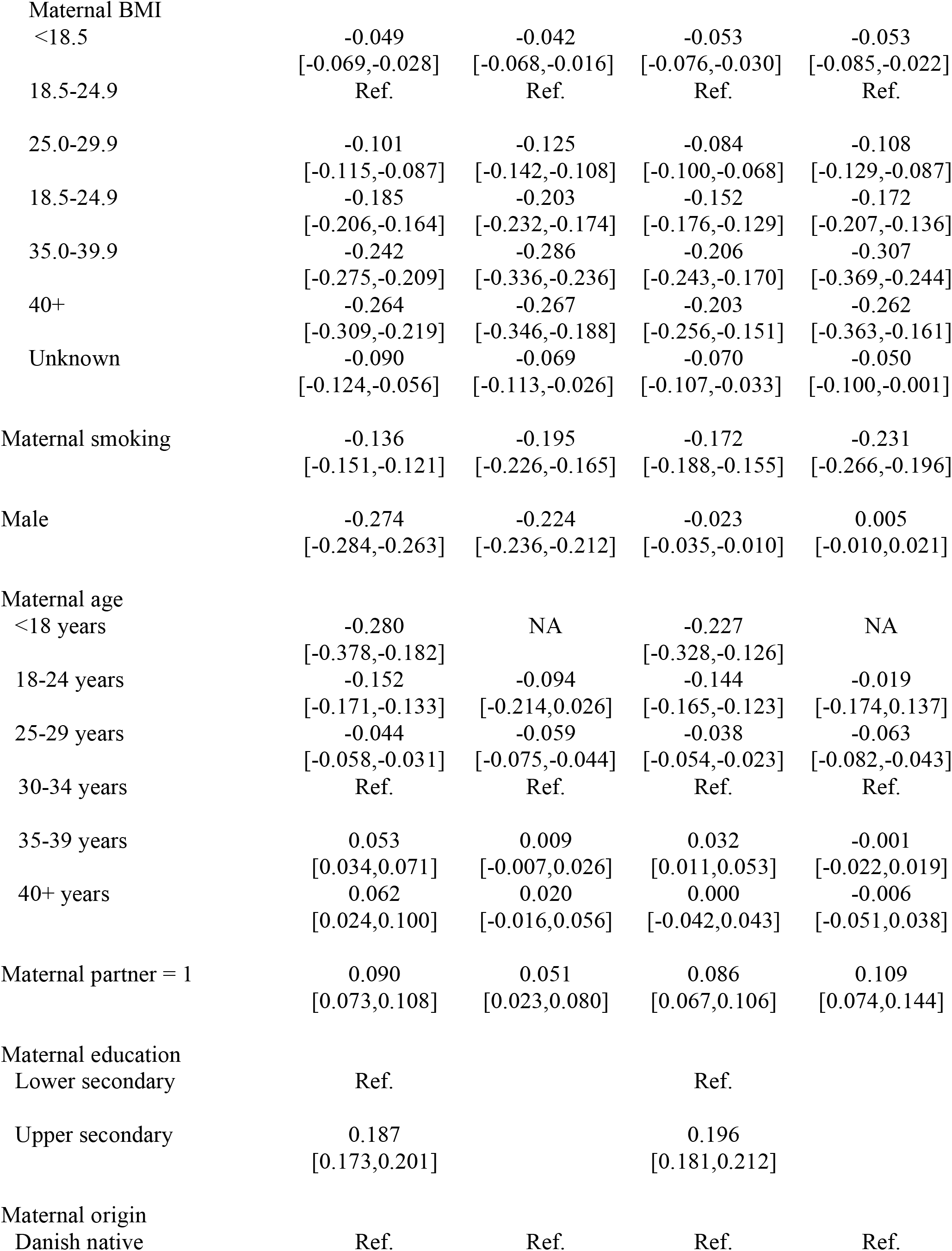
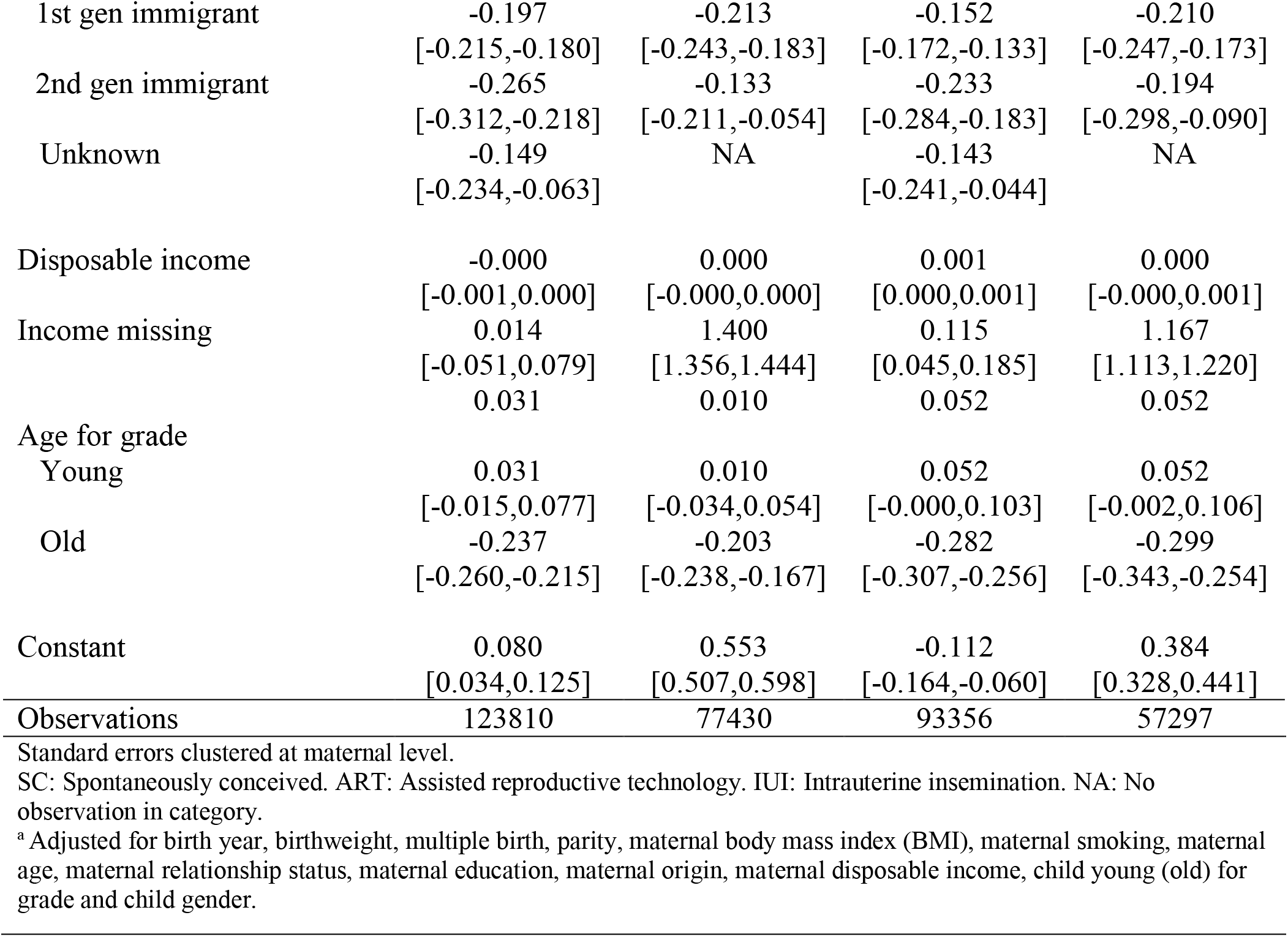
Linear model regressing second grade language comprehension and third grade math score on mode of conception and covariates by maternal education

**Table A5:**
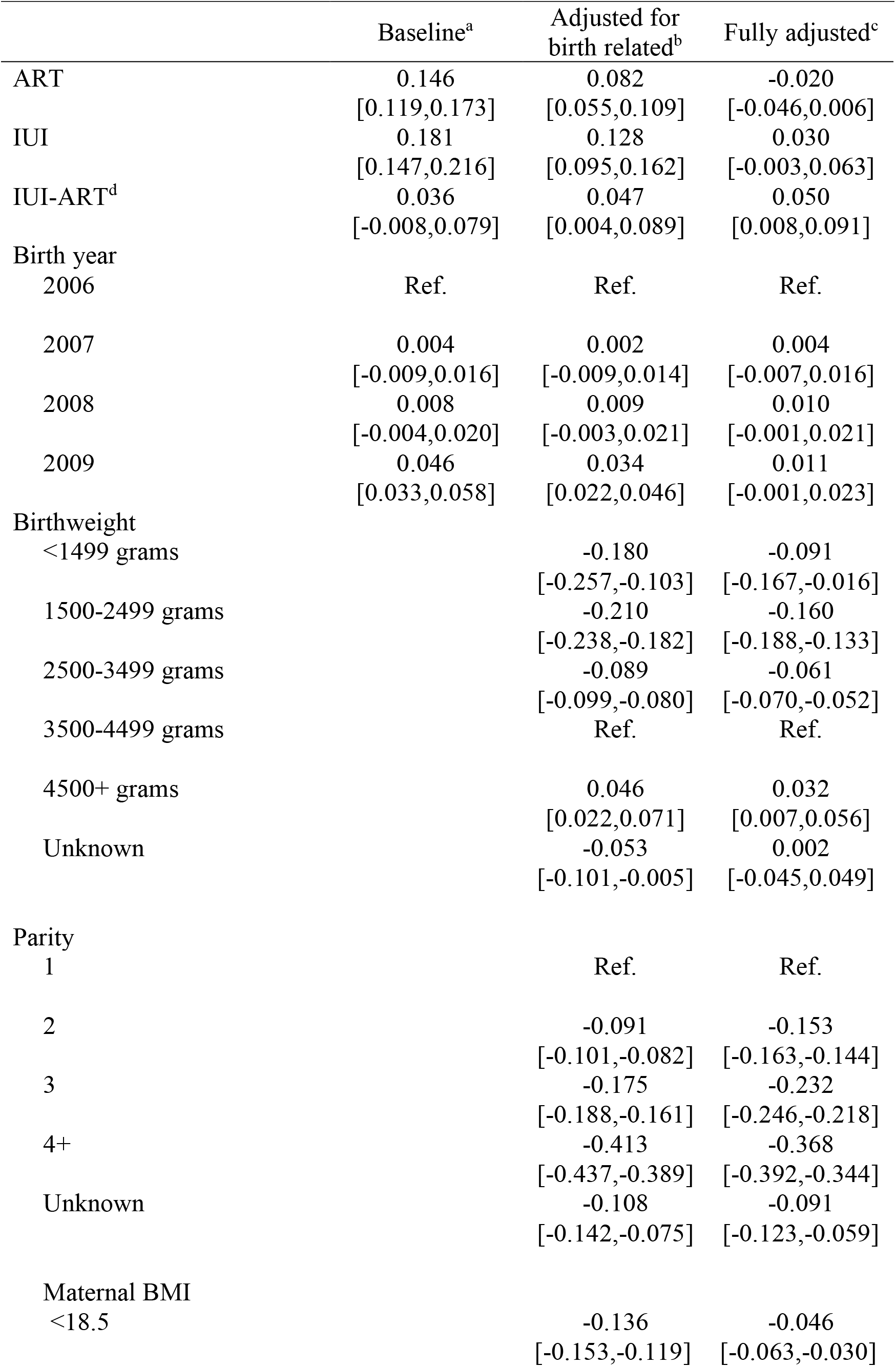
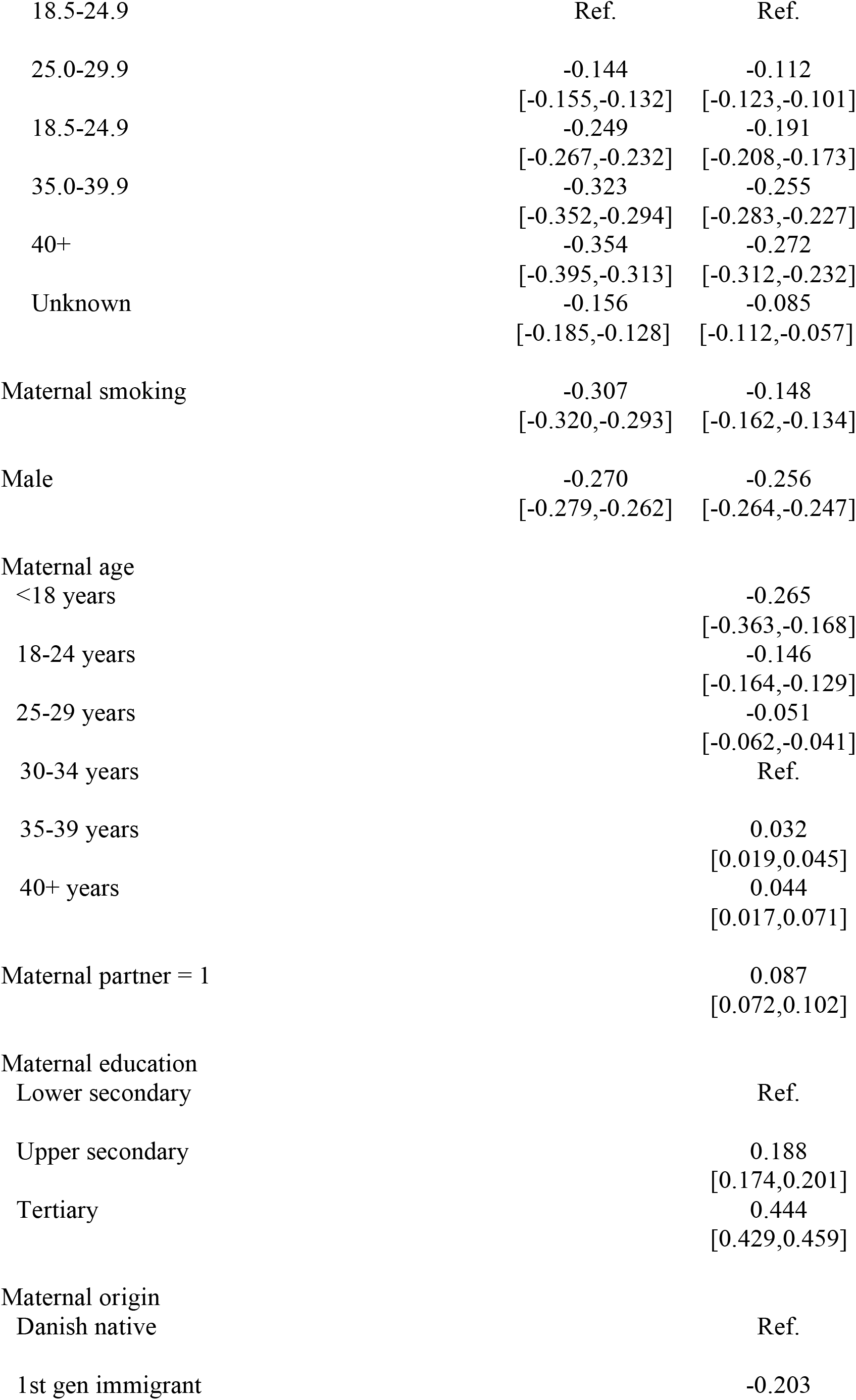
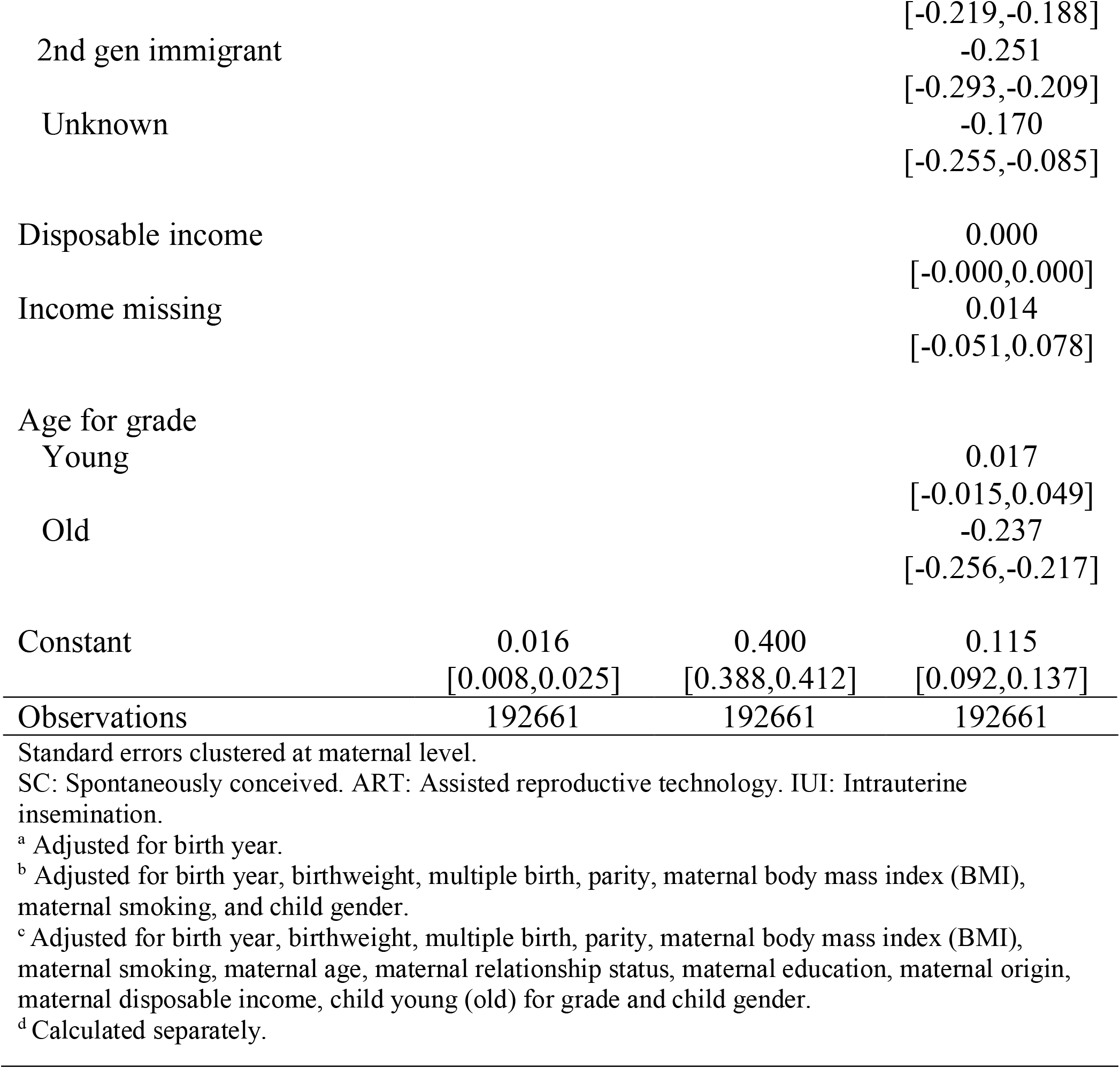
Linear model regressing second grade language comprehension score on mode of conception and covariates for singleton births

**Table A5:**
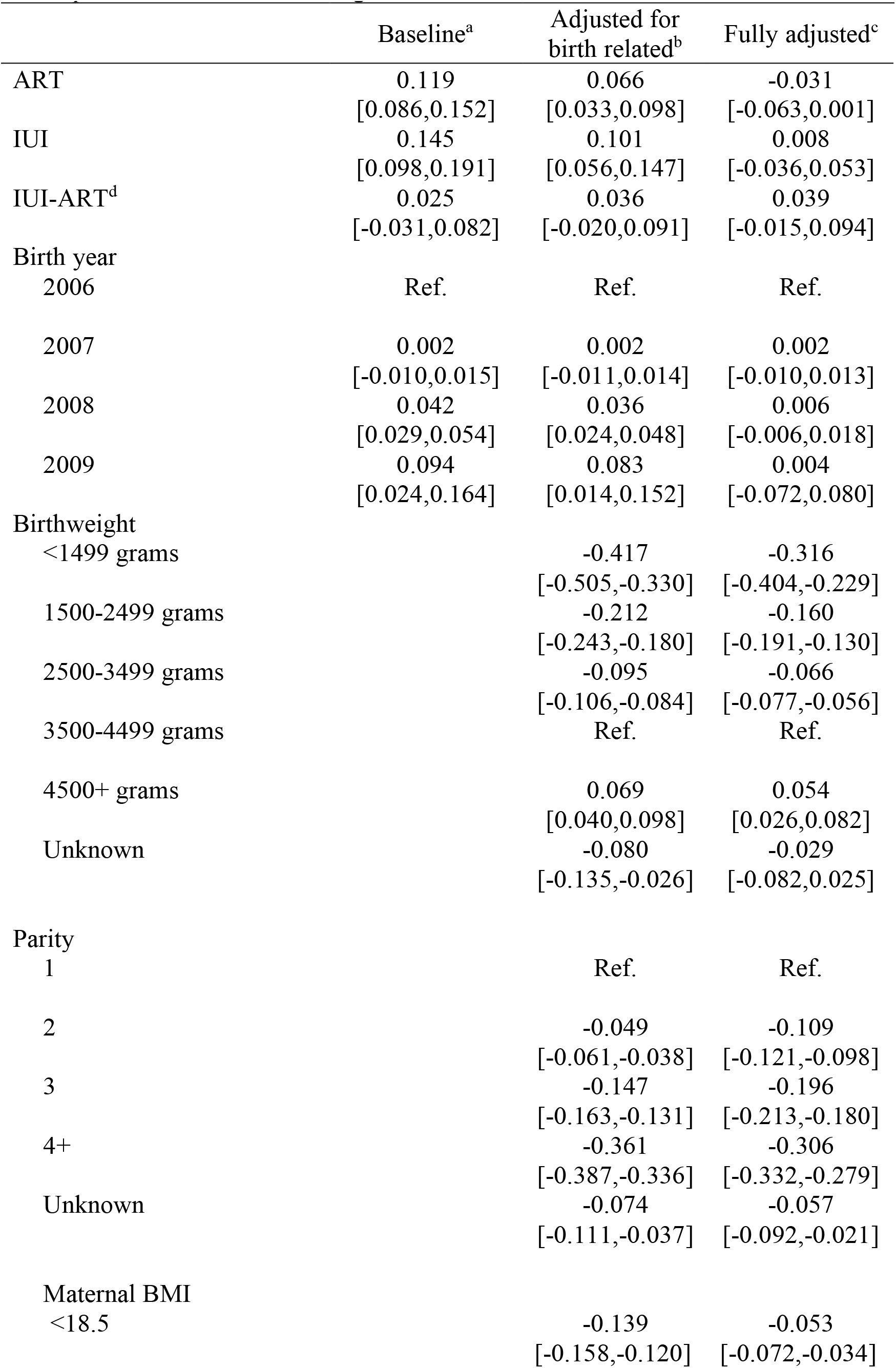
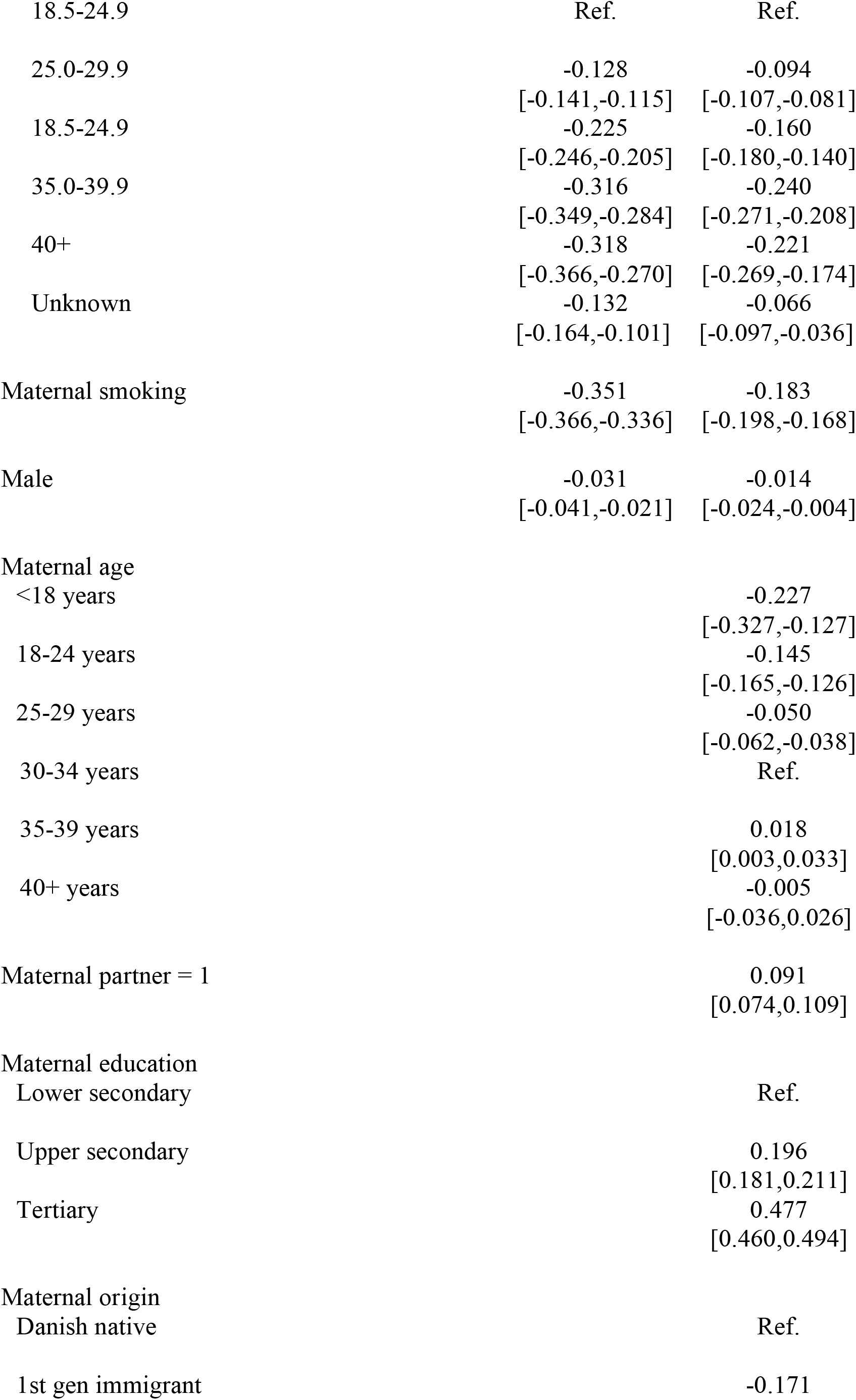
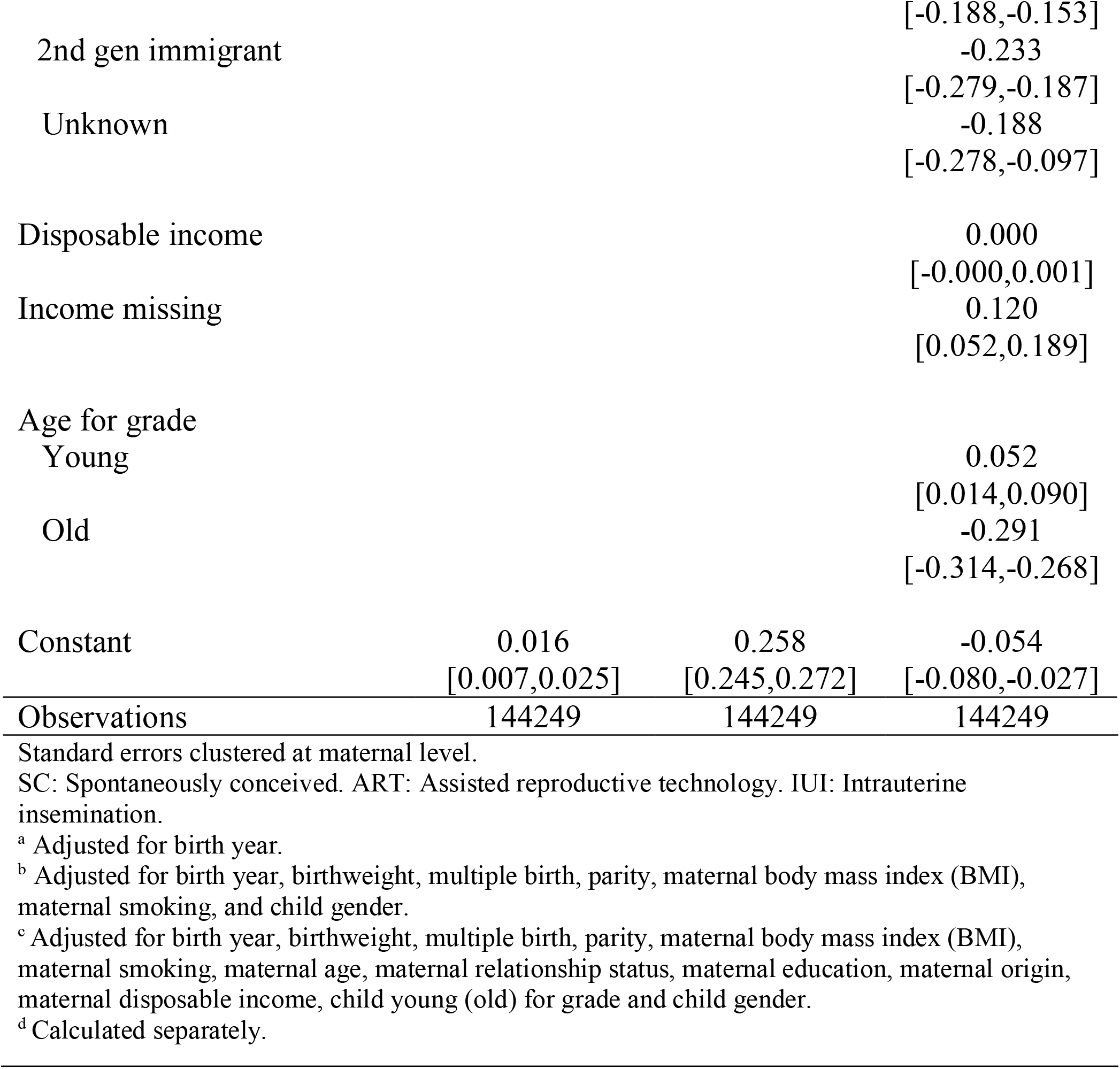
Linear model regressing second grade language comprehension score on mode of conception and covariates for singleton births

**Table A7:**
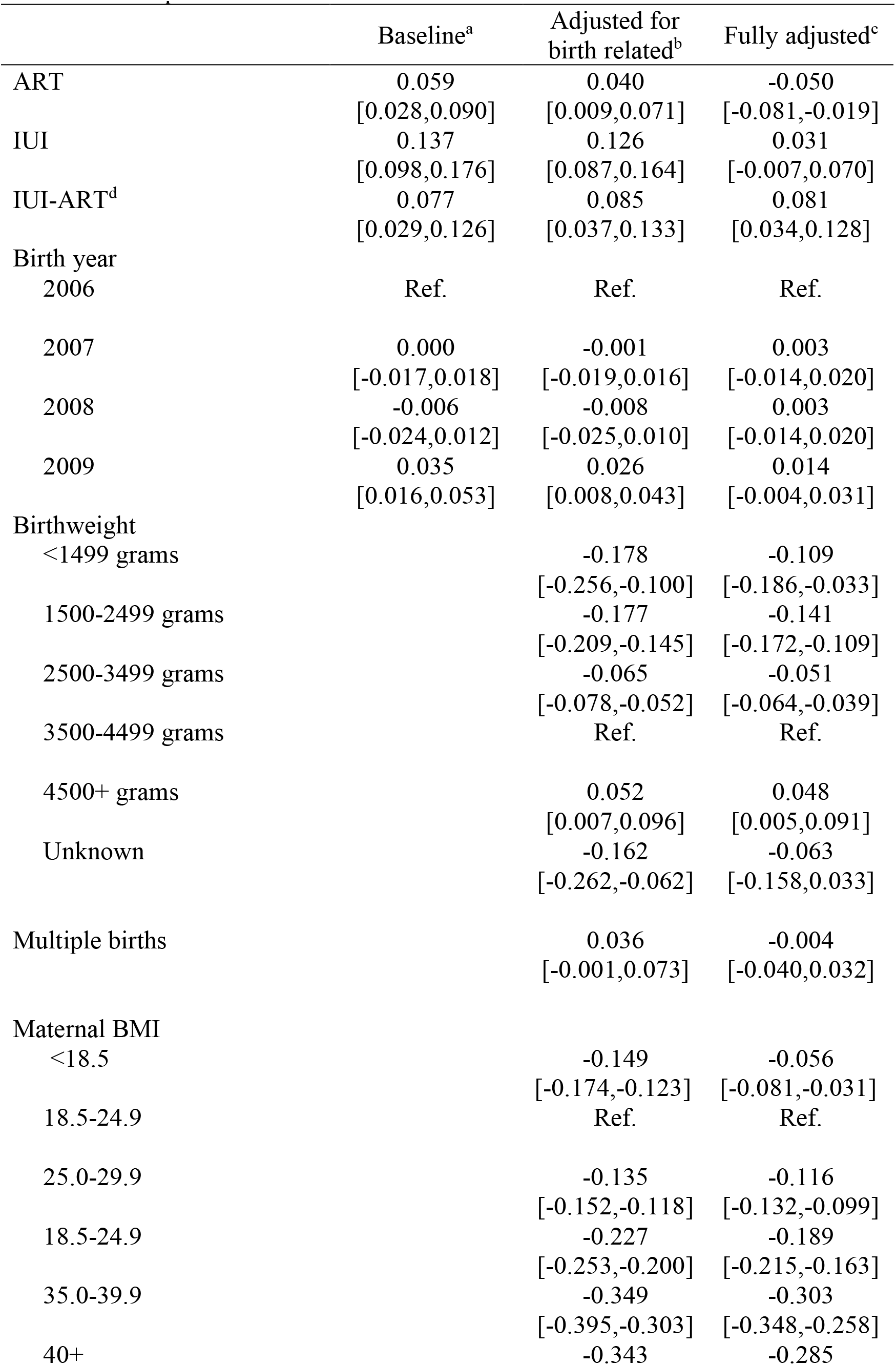
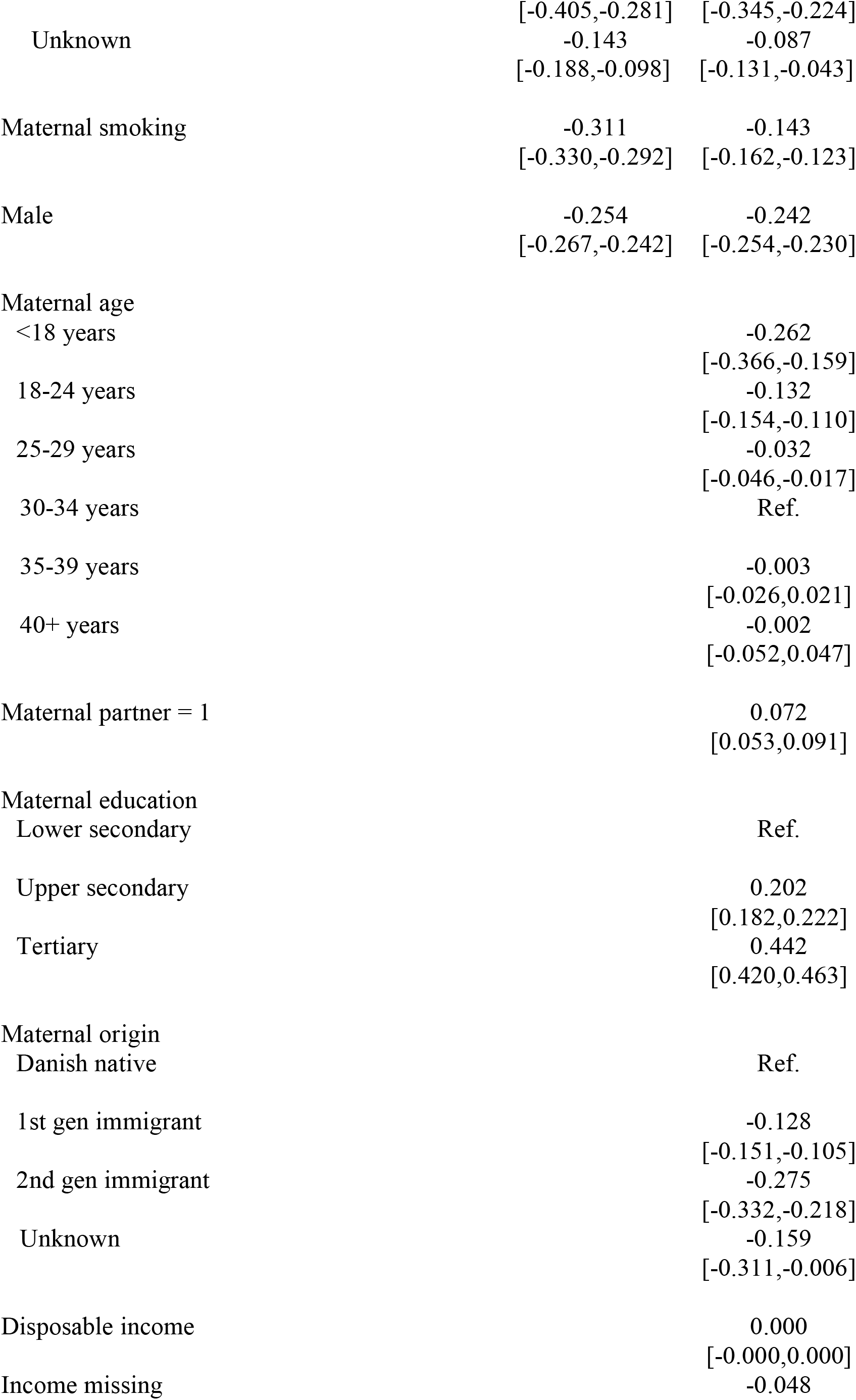
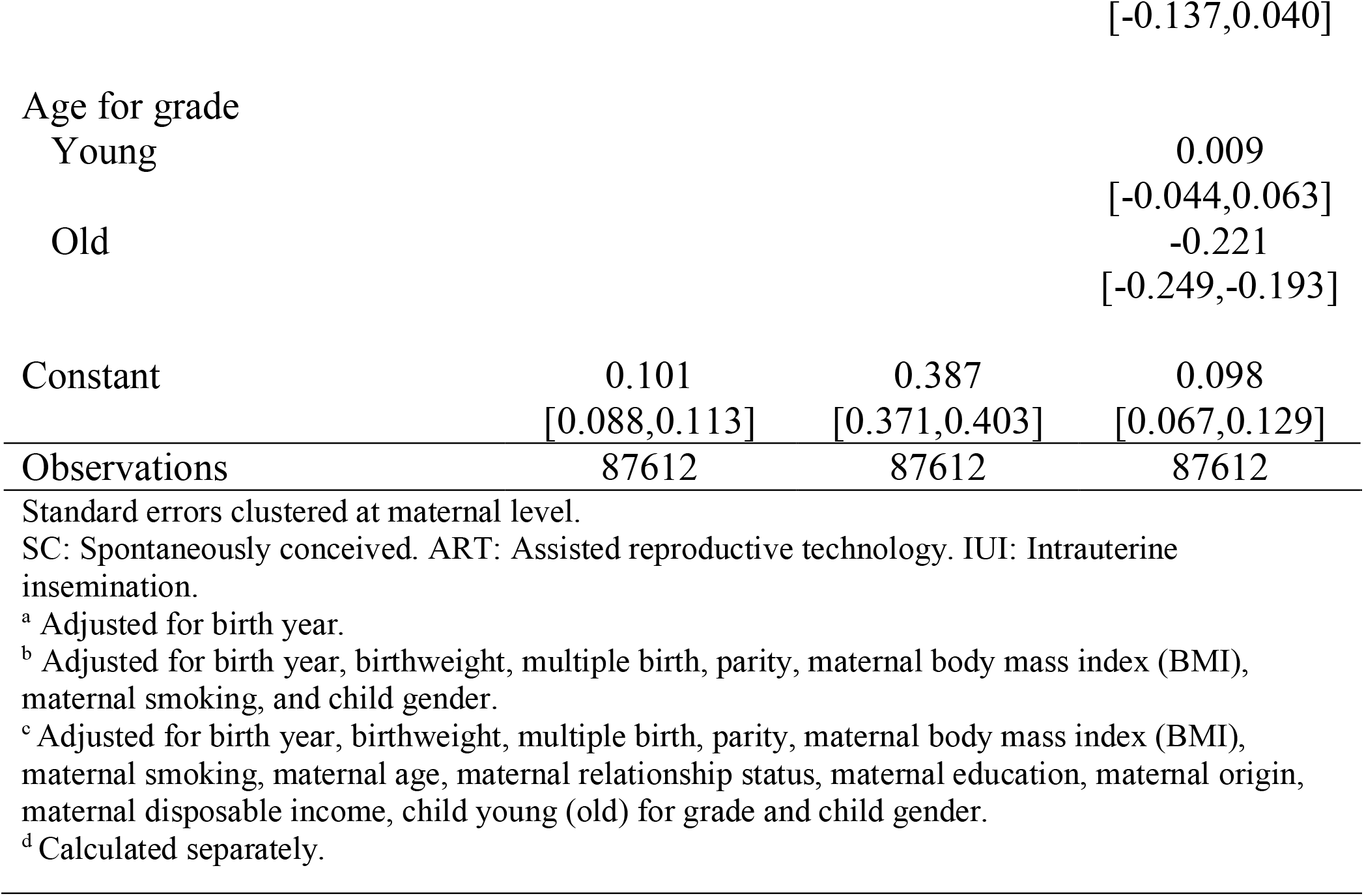
Linear model regressing second grade language comprehension score on mode of conception and covariates for first births

**Table A8:**
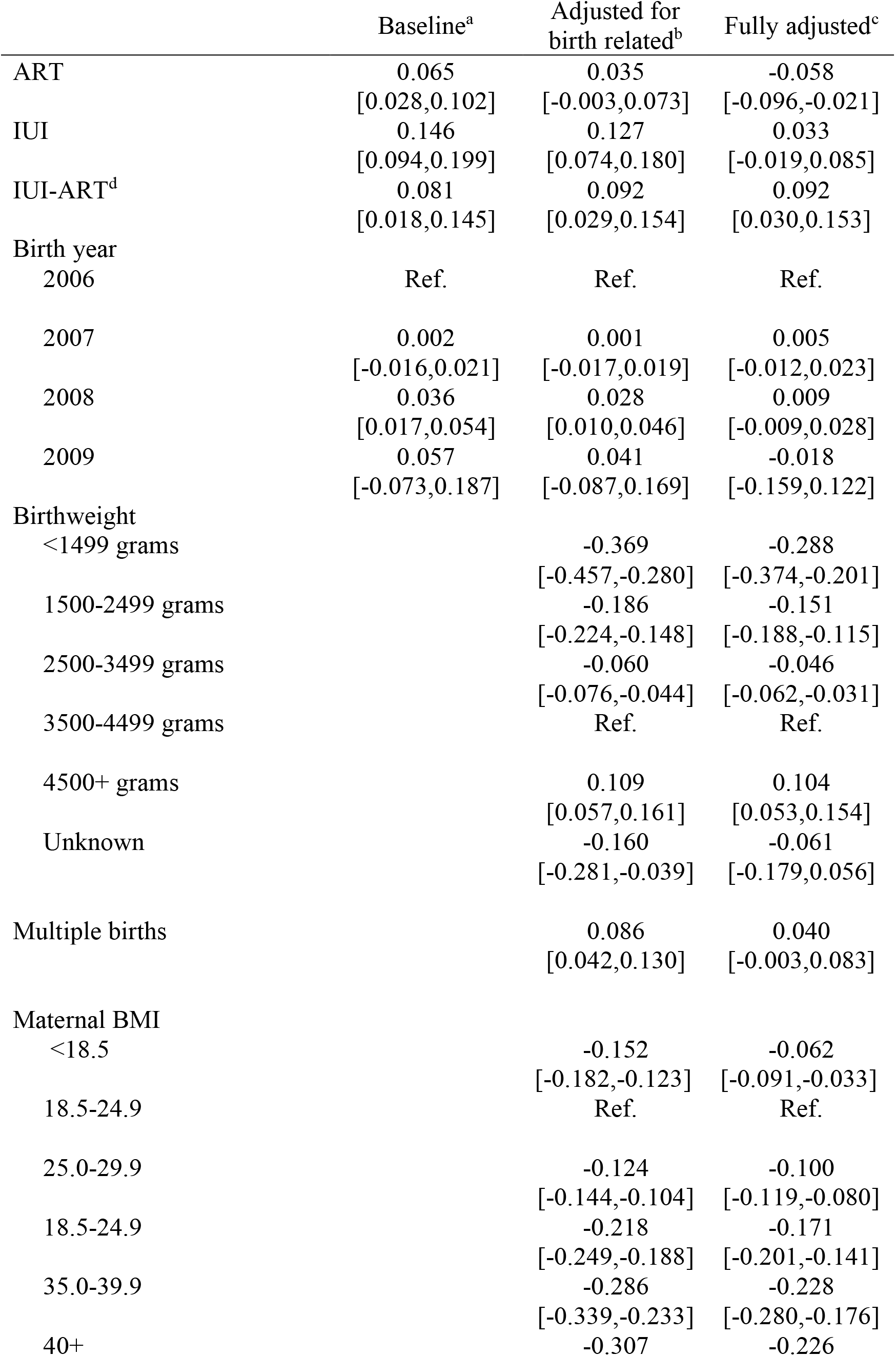
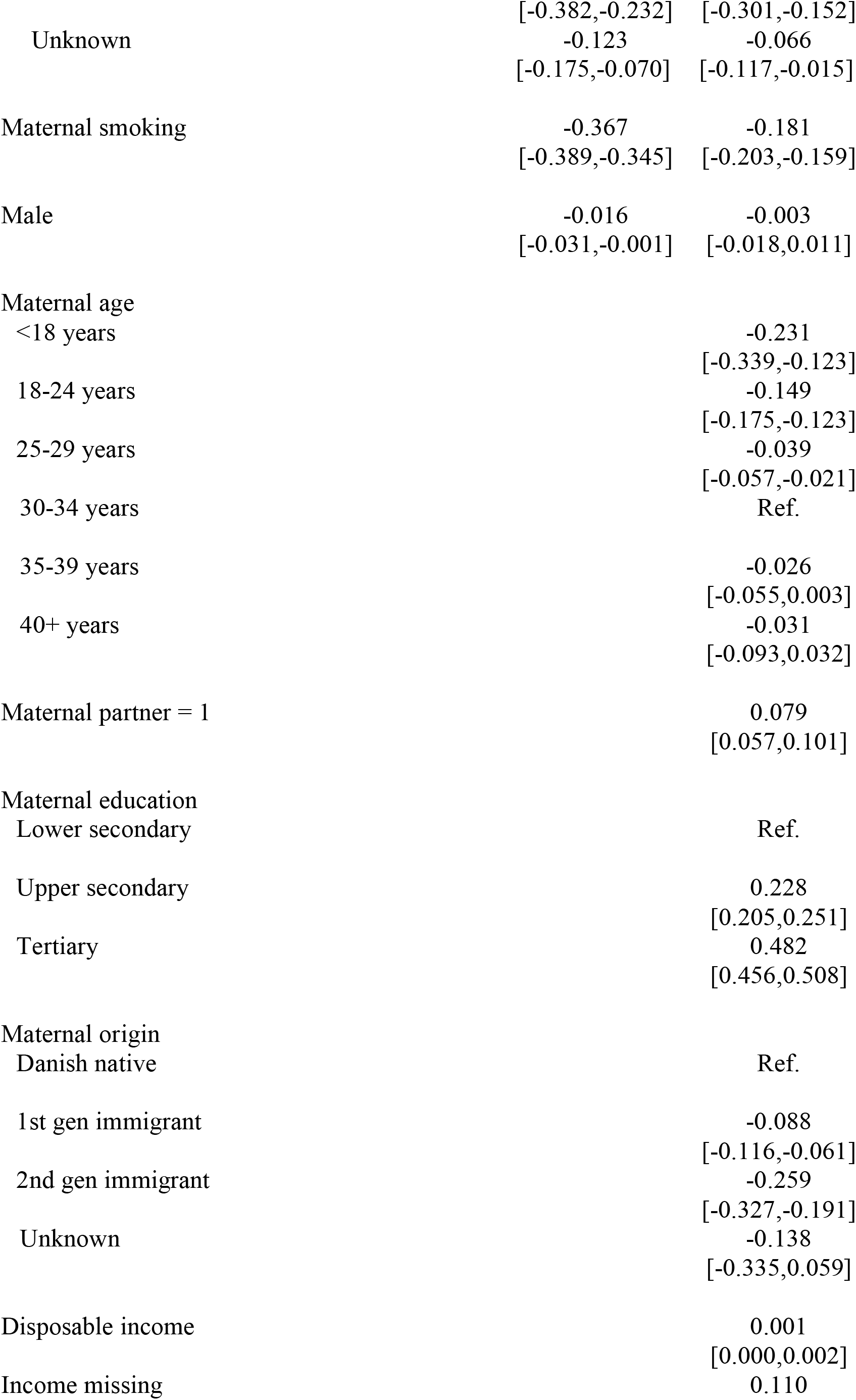
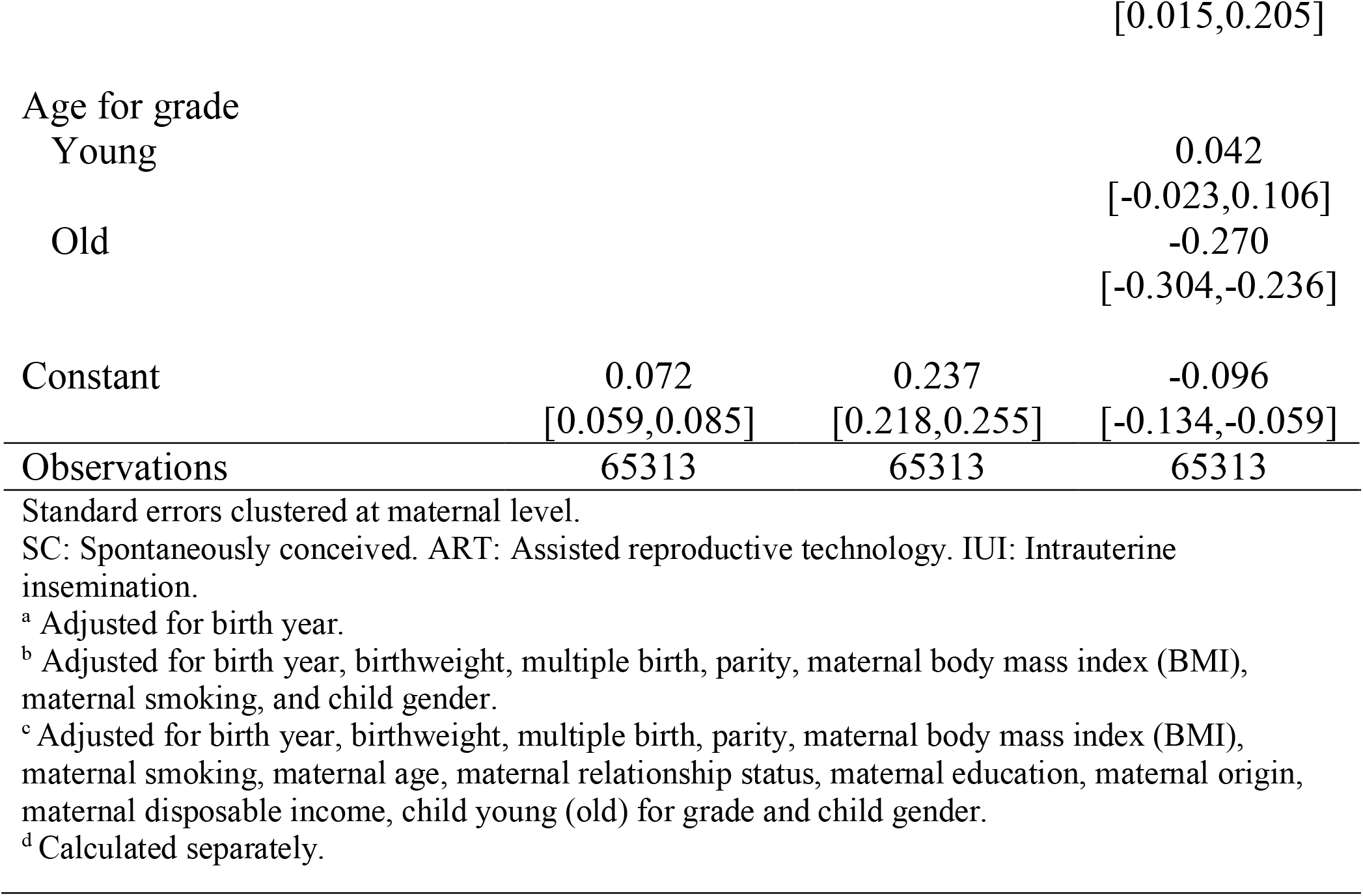
Linear model regressing third grademathematics score on mode of conception and covariates for first births

