## Supplementary material for "The association between type of conception through medically assisted reproduction and childhood cognition: A Danish population-wide cohort study": 00master.do

```
*******************************************************************************
*******************************************************************************
**	MASTER FILE
**	11-26-2022
*******************************************************************************
*******************************************************************************


global dorg "ADMIN DATA"
global data "WORK FOLDER"


cd "IVF LOCATION"
import sas using t_grund.sas7bdat, clear
cd "$data"
save IVF2.dta, replace

do 01build_sample.do

do 02build_covar.do
```
