## Supplementary material for "The association between type of conception through medically assisted reproduction and childhood cognition: A Danish population-wide cohort study": 01build_sample.do

```
forvalue t = 2006/2009{
	if `t' ==2006 use $dorg/mfr2006, clear
	else append using $dorg/mfr`t'
}

rename cpr_moder pnr
keep if pnr !=""
bysort pnr: gen help =_n

*Keep live births
keep if levende_eller_doedfoedt!="Dødfødt" 

tempfile mfr
save `mfr'

sum help
local q = r(max)
tempfil ivf
forvalue t=1/`q'{
	use `mfr' if help==`t'
	merge 1:m pnr using $data/data/IVF2.dta, keep(1 3) keepus(D_STARTDATO C_BEHANDLING V_SAMLIVS_STATUS) nogen
	
	*Remove spontanous conceptions registered in IVF registry
	replace C_B ="" if inlist(C_B,"SPONTAN")
	gen diff = foedselsdato-D_START
	
	*Mark treatment occurinf within 10 months
	gen ivf =  diff < 365.2425*10/12 & diff > 0
	bysort pnr (ivf): keep if _n==_N
	if `t' > 1 append using `ivf'
	
	save `ivf', replace
	
}

gen IUI = C_B =="IUI" & ivf==1
gen ART = !inlist(C_B,"","SPONTAN") & IUI ==0 & ivf==1
replace C_B = "" if ivf==0
replace C_B = "OOCYT" if inlist(C_B,"OOCYT_DONOR","DISTRI")
replace C_B = "FER" if C_B == "ALM+FER"
replace C_B = "IVF/ICSI" if C_B =="ALM"
drop D_START

save $data/data/ivf_sample, replace
```
