## Supplementary material for "The association between type of conception through medically assisted reproduction and childhood cognition: A Danish population-wide cohort study": 02build_covar.do

```
	tempfile covar

		
	forvalue t =2005/2008{
		local n = `t'-1
		local m = `t'+1
		use $data/data/ivf_sample if year(foedselsdato)==`m', clear
		destring pnr, replace force
		keep if pnr !=.
		*Education captured end of year prior to birth
		merge m:1 pnr using e:/data/rawdata/705830/udda`t', ///
			keepus(hfaudd) keep(1 3) nogen
		*BEF info brought in prior year
		merge m:1 pnr using e:/data/rawdata/705830/bef12_`t', ///
			keepus(ie_type e_faelle_id civst antpersh antboernh) keep(1 3) nogen
		rename ie_type IE_TYPE
		*Widen scope to capture immigration background
		merge m:1 pnr using e:/data/rawdata/705830/bef12_`n', ///
			keepus(ie_type ) keep(1 3) nogen
		replace IE_TYPE = ie_type if IE_TYPE ==.
		drop ie_type
		merge m:1 pnr using e:/data/rawdata/705830/bef12_`m', ///
			keepus( ie_type ) keep(1 3) nogen
		replace IE_TYPE = ie_type if IE_TYPE ==.
		drop ie_type
		rename IE_TYPE ie_type
		**Obtain income from year proir to birth
		merge m:m  pnr using e:/data/rawdata/705830/ind`t', ///a few income observations have double 		observations, but does not affaect dispon_13
			keepus(dispon_13) keep(1 3) nogen 
		bysort cpr_barn: keep if _n ==1 // correct for m:m merge at income
		cap rename *`t' *
		
		if `t' > 2005 append using `covar'
		save `covar', replace
	}


	
	gen start=string(hfaudd,"%04.0f")
	merge m:1 start using "\\SRVFSENAS1\data\Formater\SAS formater i Danmarks Statistik\STATA_datasaet\Disced\c_audd_level_l1l4_k.dta" , keep(1 3) nogen
	 
	destring AUDD, gen(isced)
	drop AUDD start
	
	**ISCED code education, missing coded as lowest level
	gen level = 1 if isced < 3 | isced ==. | isced ==9
	replace level = 2 if inlist(isced,3,4)
	replace level = 3 if level ==.
	
	rename dispon_13 disposable_income
	
	**Partner if cohabiting, married/partnership or residing in a hosuehold with one other adult
	gen partner = e_faelle_id !="" | civst==("G","P") | (antpersh-antboernh ==2)
	
	 
	save $data/data/ivf_covar.dta, replace
```
