## Supplementary material for "The association between type of conception through medically assisted reproduction and childhood cognition: A Danish population-wide cohort study": 03build_testdata.do

```
*******************************************************************
*******************************************************************
*******************************************************************
*******************************************************************
****
****	build_outcomes.do - generate subject specific standardized
****	test (z_score) for math and language
****	
****	
****			11-26-2022
****	
****
****
****
	* pnr_child - child identifier
	* subject - test subject
	* testtime - time and date of test
	* school_id - school identifier
	* grade - grade level
	* z_score - standardized grade
	

*******************************************************************
*******************************************************************
*******************************************************************


use "E:\Data\workdata\705830\JH\data\nationale_testdata.dta"  ///
	if testtype =="obligatorisk", clear

	
	**Keep relevant tests

	replace klassetrin =0 if klassetrin ==.
	bysort pnr skoleaar: egen kl = max(klassetrin)		
	gen disqualify	=1
		foreach x in 2 3 4 6 8{
		replace disqualify = 0 if kl ==`x' & !regexm(fag,`"`x'"')
	}
	
	gen subject_matter =""
	foreach x in Biologi Dansk/læsning Engelsk Fysik/kemi Geografi Matematik{
	    replace subject_matter = "`x'" if regexm(fag,"`x'")
	}

	gen subject_grade = .
	forvalue t=2/8{
	    replace subject_grade= `t' if regexm(fag,"`t'")
	}
	
	drop if disqualify ==0
	
	bysort pnr: gen help =_N

	tab help 
	
	bysort pnr skoleaar: gen help1 =_N

	tab help1
	
	
	**keep relevant variables
	keep pnr skoleaar fag klassetrin testtid theta* instnr  subject_matter subject_grade
	
	destring pnr, replace force
	drop if pnr ==.
	
	**standardize variables from logit scale across domains
	foreach x in 1 2 3{
		bysort subject_matter subject_grade skoleaar: egen mean = mean(theta_p`x')
		bysort subject_matter subject_grade skoleaar: egen   sd = sd(theta_p`x')
		gen ztheta`x' = (theta_p`x'-mean)/sd
		drop sd mean
	}
	
	*generate joint score
	gen z_prelim = (ztheta1+ztheta2+ztheta3)/3
	bysort skoleaar fag: egen mean = mean(z_prelim)
	bysort skoleaar fag: egen sd   = sd(z_prelim)
	gen z_score = (z_prelim-mean)/sd
	drop z_prelim mean sd
	
	gen year = substr(testtid,1,4)
	gen month= substr(testtid,6,2)
	gen day  = substr(testtid,9,2)
	
	
	destring year, replace
	destring month, replace
	destring day, replace
	
	
	drop theta* ztheta*

	rename pnr pnr_child
	rename fag subject
	rename testtid testtime
	rename instnr school_id
	rename klassetrin grade
	
	save $data\test_data.dta, replace
	
```
