## Supplementary material for "The association between type of conception through medically assisted reproduction and childhood cognition: A Danish population-wide cohort study": 04estimate.do

```
**find migration

use pnr handdto indud using "$dorg/vnds2021.dta", clear

bysort pnr (hand): keep if _n==_N
keep if indud==2
drop indud
tempfile migrant
save `migrant'

use $data/data/ivf_covar.dta, clear

rename pnr mor_id
destring cpr_barn, gen(pnr_child) force
keep if pnr_child !=.

merge 1:m pnr_child using $data\data\test_data.dta, keep(1 3)

keep if subject_grade < 4 | subject_grade==. 

gen byear = year(foedselsdato)

**remove children who migrate/die before age 9
rename pnr_child pnr
merge m:1 pnr using $dorg/dod2020, keep(1 3) nogen
drop if alder_haend < 9
merge m:1 pnr using `migrant', keep(1 3) nogen
keep if (handdto-foedselsdato)/365.2425 >=9

*recode data

gen twin_plus =  flerfoldsfoedsel_beregnet >1
label var twin_plus "Multiple birth"

replace vaegt_b =. if vaegt_b < 450 | vaegt_b ==9999
recode vaegt_b (450/1499=1 "450-1499 grams") (1500/2499=2 "1500-2499 grams") ///
			(2500/3499=3 "2500-3499 grams")  (3500/4499=4 "3500-4499 grams") ///
			(4500/9000=5 "4500+ grams") (.=6 "Unknown")  , gen(weight)
label var weight "Birthweight"
recode alder_moder 	(0/17 =1 "<18 years") (18/24 =2 "18-24 years") ///
					(25/29 = 3 "25-29 years") (30/34 = 4 "30-34 years") ///
					(35/39 = 5 "35-39 years") (40/61 = 6 "40+ years"), ///
					gen(maternal_age)
label var maternal_age "Maternal age"

destring paritet, replace force 
recode paritet 	(. = 5 "Unknown") (1=1 "Parity 1") (2=2 "2") (3=3 "3") (4/. = 4 "4+"), ///
				gen(parity)
label var parity "Parity"				
		
destring ryg, replace force
recode ryg (1/98 =1 "Smoker") (else = 0 "Non-smoker"), gen(smoker)
label var smoker "Smoked during pregnancy"

label var parity "Parity"

*order parity
bysort mor_id byear: egen helpX= min(parity)
replace parity = helpX if helpX != parity
drop helpX

**correct error in BMI coding
replace bmi_moder = bmi_moder/10 if bmi_moder > 100
replace vaegt_moder = vaegt_moder /100 if vaegt_moder > 3000
replace vaegt_moder = vaegt_moder /10 if vaegt_moder > 1000
replace hoejde_moder = hoejde_moder /10 if hoejde_moder > 1000
replace hoejde_moder = hoejde_moder +100 if hoejde_moder < 100
gen bmi2= vaegt_moder/((hoejde_moder/100)^2)
replace bmi_moder = bmi2 if bmi2 < bmi_moder & bmi2 < 55


recode bmi_moder 	(0/18.499 = 1 " 18.5") (18.5/24.999 = 2 "18.5-24.9") ///
					(25/29.999 = 3 "25.0-29.9") (30.0/34.999 = 4 "30.0-34.9") ///
					(35/39.999 = 5 "35.0-39.9") (40/55 = 6 "40+") ///
					(0 55.001/99 . = 99 "Unknown"), gen(maternal_bmi)
label var maternal_bmi "Maternal BMI"
					
rename ie_ty ie
recode ie (1=1 "Native Dane") (2=2 "1st G Migrant") (3=3 "2nd G Migrant") (.=4 "Unknown"), gen(ie_type)
drop ie
label var ie_type "Migration background"

gen disp_mis = dispo ==.
label var disp_mis "Income missing"
replace dispo = 0 if dispo ==.	
replace dispo = dispo/7450
label var dispo "Disp income in €1K"	

label var level "Maternal level of Education"

gen young = year-byear <= 6+subject_grade
label var young "Young for grade"
gen old = year-byear > 6+subject_grade+1
label var young "Old for grade"
gen male = koen=="M"
label var male "Male"
label var ivf "MAR"
label var byear "Birth year"
label var partner "In relatoinship"

**IS IVF correlated with not in sample

bysort pnr: gen mark= _n ==1
gen in_sample = (z_score !=. & subject_grade < 4) if mark==1
drop mark

**ABSENT FROM OUTCOME

reg in_sample i.ivf i.byear
eststo e1
margins i.ivf
reg in_sample i.ivf i.byear i.b4.weight twin_plus i.b4.maternal_age i.parity i.b2.maternal_bmi smoker partner i.level i.ie_type dispo disp_  male
eststo e2
margins ivf

esttab e* using $data/tables/TableA1.rtf, plain par b(3) ci(3) label replace


drop if subject_grade ==.

gen TREATED = "NC"
replace TREATED = "ART" if ART==1
replace TREATED = "IUI" if IUI==1

tab subject_grade TREATED, col
tab byear TREATED, col
tab weight TREATED, col
tab twin_plus TREATED, col
tab maternal_age TREATED, col
tab parity TREATED, col
tab maternal_bmi TREATED, col 
tab smoker TREATED, col 
tab partner TREATED, col 
tab level TREATED, col 
tab ie_type TREATED, col 
tab young TREATED, col
tab old TREATED, col
tab male TREATED, col
table () (TREATED), stat(mean dispo) stat(sd dispo), if subject_grade==2

encode TREATED, gen(treated)

reg z_score i.byear i.b3.treated if subject_grade==2, cl(mor_id)
eststo e1
margins treated
reg z_score i.byear i.b3.treated i.b4.weight twin_plus i.parity i.b2.maternal_bmi smoker male if subject_grade==2, cl(mor_id)
eststo e2
margins treated
reg z_score i.byear i.b3.treated i.b4.weight twin_plus i.b4.maternal_age i.parity i.b2.maternal_bmi smoker partner i.level i.ie_type dispo disp_ young old male if subject_grade==2, cl(mor_id)
eststo e3
margins treated

esttab e* using $data/tables/grade2_both.rtf, plain par b(3) ci(3) label replace


reg z_score i.byear i.b3.treated if subject_grade==3, cl(mor_id)
eststo e1
margins treated
reg z_score i.byear i.b3.treated i.b4.weight twin_plus i.parity i.b2.maternal_bmi smoker male  if subject_grade==3, cl(mor_id)
eststo e2
margins treated
reg z_score i.byear i.b3.treated i.b4.weight twin_plus i.b4.maternal_age i.parity i.b2.maternal_bmi smoker partner i.level i.ie_type dispo disp_ young old male if subject_grade==3, cl(mor_id)
eststo e3
margins treated
esttab e* using $data/tables/grade3_both.rtf, plain par b(3) ci(3) label replace

gen NC = ART==0 & IUI ==0

reg z_score i.byear NC IUI if subject_grade==2, cl(mor_id)
eststo e1
reg z_score i.byear NC IUI  i.b4.weight twin_plus i.parity i.b2.maternal_bmi smoker male   if subject_grade==2, cl(mor_id)
eststo e2
reg z_score i.byear NC IUI i.b4.weight twin_plus i.b4.maternal_age i.parity i.b2.maternal_bmi smoker partner i.level i.ie_type dispo disp_ young old male if subject_grade==2, cl(mor_id)
eststo e3

esttab e* using $data/tables/grade2_IUI.rtf, plain par b(3) ci(3) label replace
esttab e*, keep(IUI) ci(3) b(3)

reg z_score i.byear NC IUI if subject_grade==3, cl(mor_id)
eststo e1
reg z_score i.byear NC IUI  i.b4.weight twin_plus i.parity i.b2.maternal_bmi smoker male   if subject_grade==3, cl(mor_id)
eststo e2
reg z_score i.byear NC IUI i.b4.weight twin_plus i.b4.maternal_age i.parity i.b2.maternal_bmi smoker partner i.level i.ie_type dispo disp_ young old male if subject_grade==3, cl(mor_id)
eststo e3

esttab e* using $data/tables/grade3_IUI.rtf, plain par b(3) ci(3) label replace


********************************************************************************
********************************************************************************
**		PARITY 1
**
********************************************************************************
********************************************************************************
snapshot erase _all
snapshot save
keep if parity ==1

reg z_score i.byear i.b3.treated if subject_grade==2, cl(mor_id)
eststo e1
margins treated
reg z_score i.byear i.b3.treated i.b4.weight twin_plus i.b2.maternal_bmi smoker male if subject_grade==2, cl(mor_id)
eststo e2
margins treated
reg z_score i.byear i.b3.treated i.b4.weight twin_plus i.b4.maternal_age i.b2.maternal_bmi smoker partner i.level i.ie_type dispo disp_ young old male if subject_grade==2, cl(mor_id)
eststo e3
margins treated

esttab e* using $data/tables/grade2_both_par1.rtf, plain par b(3) ci(3) label replace


reg z_score i.byear i.b3.treated if subject_grade==3, cl(mor_id)
eststo e1
margins treated
reg z_score i.byear i.b3.treated i.b4.weight twin_plus i.b2.maternal_bmi smoker male  if subject_grade==3, cl(mor_id)
eststo e2
margins treated
reg z_score i.byear i.b3.treated i.b4.weight twin_plus i.b4.maternal_age i.b2.maternal_bmi smoker partner i.level i.ie_type dispo disp_ young old male if subject_grade==3, cl(mor_id)
eststo e3
margins treated
esttab e* using $data/tables/grade3_both_par1.rtf, plain par b(3) ci(3) label replace

reg z_score i.byear NC IUI if subject_grade==2, cl(mor_id)
eststo e1
reg z_score i.byear NC IUI  i.b4.weight twin_plus i.b2.maternal_bmi smoker male   if subject_grade==2, cl(mor_id)
eststo e2
reg z_score i.byear NC IUI i.b4.weight twin_plus i.b4.maternal_age i.b2.maternal_bmi smoker partner i.level i.ie_type dispo disp_ young old male if subject_grade==2, cl(mor_id)
eststo e3

esttab e* using $data/tables/grade2_IUI_par1.rtf, plain par b(3) ci(3) label replace
esttab e*, keep(IUI) ci(3) b(3)

reg z_score i.byear NC IUI if subject_grade==3, cl(mor_id)
eststo e1
reg z_score i.byear NC IUI  i.b4.weight twin_plus i.parity i.b2.maternal_bmi smoker male   if subject_grade==3, cl(mor_id)
eststo e2
reg z_score i.byear NC IUI i.b4.weight twin_plus i.b4.maternal_age i.parity i.b2.maternal_bmi smoker partner i.level i.ie_type dispo disp_ young old male if subject_grade==3, cl(mor_id)
eststo e3

esttab e* using $data/tables/grade3_IUI_par1.rtf, plain par b(3) ci(3) label replace


*******************************************************************************
*******************************************************************************
**	Keep singleton birth
*******************************************************************************
*******************************************************************************

snapshot use 1

keep if twin_ ==0
reg z_score i.byear i.b3.treated if subject_grade==2, cl(mor_id)
eststo e1
margins treated
reg z_score i.byear i.b3.treated i.b4.weight i.parity  twin_plus i.b2.maternal_bmi smoker male if subject_grade==2, cl(mor_id)
eststo e2
margins treated
reg z_score i.byear i.b3.treated i.b4.weight i.parity  twin_plus i.b4.maternal_age i.b2.maternal_bmi smoker partner i.level i.ie_type dispo disp_ young old male if subject_grade==2, cl(mor_id)
eststo e3
margins treated

esttab e* using $data/tables/grade2_both_singleton1.rtf, plain par b(3) ci(3) label replace


reg z_score i.byear i.b3.treated if subject_grade==3, cl(mor_id)
eststo e1
margins treated
reg z_score i.byear i.b3.treated i.b4.weight i.parity  twin_plus i.b2.maternal_bmi smoker male  if subject_grade==3, cl(mor_id)
eststo e2
margins treated
reg z_score i.byear i.b3.treated i.b4.weight i.parity  twin_plus i.b4.maternal_age i.b2.maternal_bmi smoker partner i.level i.ie_type dispo disp_ young old male if subject_grade==3, cl(mor_id)
eststo e3
margins treated
esttab e* using $data/tables/grade3_both_singleton1.rtf, plain par b(3) ci(3) label replace

reg z_score i.byear NC IUI if subject_grade==2, cl(mor_id)
eststo e1
reg z_score i.byear NC IUI  i.b4.weight i.parity  twin_plus i.b2.maternal_bmi smoker male   if subject_grade==2, cl(mor_id)
eststo e2
reg z_score i.byear NC IUI i.b4.weight i.parity  twin_plus i.b4.maternal_age i.b2.maternal_bmi smoker partner i.level i.ie_type dispo disp_ young old male if subject_grade==2, cl(mor_id)
eststo e3

esttab e* using $data/tables/grade2_IUI_singleton1.rtf, plain par b(3) ci(3) label replace
esttab e*, keep(IUI) ci(3) b(3)

reg z_score i.byear NC IUI if subject_grade==3, cl(mor_id)
eststo e1
reg z_score i.byear NC IUI  i.b4.weight twin_plus i.parity i.b2.maternal_bmi smoker male   if subject_grade==3, cl(mor_id)
eststo e2
reg z_score i.byear NC IUI i.b4.weight i.parity  twin_plus i.b4.maternal_age i.parity i.b2.maternal_bmi smoker partner i.level i.ie_type dispo disp_ young old male if subject_grade==3, cl(mor_id)
eststo e3

esttab e* using $data/tables/grade3_IUI_singleton1.rtf, plain par b(3) ci(3) label replace 

********************************************************************************
**	By education
**
********************************************************************************


snapshot use 1


reg z_score i.byear i.treated i.b4.weight twin_plus i.b4.maternal_age i.parity i.b2.maternal_bmi smoker partner i.ie_type i.level dispo disp_ young old male if subject_grade==2 & level <= 2, cl(mor_id)
eststo e1
margins treated
reg z_score i.byear NC IUI i.b4.weight twin_plus i.b4.maternal_age i.parity i.b2.maternal_bmi smoker partner i.ie_type i.level dispo disp_ young old male if subject_grade==2 & level <= 2, cl(mor_id)

reg z_score i.byear i.treated i.b4.weight twin_plus i.b4.maternal_age i.parity i.b2.maternal_bmi smoker partner i.ie_type dispo disp_ young old male if subject_grade==2 & level > 2, cl(mor_id)
eststo e2
margins treated
reg z_score i.byear NC IUI i.b4.weight twin_plus i.b4.maternal_age i.parity i.b2.maternal_bmi smoker partner i.ie_type dispo disp_ young old male if subject_grade==2 & level > 2, cl(mor_id)


reg z_score i.byear i.treated i.b4.weight twin_plus i.b4.maternal_age i.parity i.b2.maternal_bmi smoker partner i.ie_type i.level dispo disp_ young old male if subject_grade==3 & level <= 2, cl(mor_id)
eststo e3
margins treated
reg z_score i.byear NC IUI i.b4.weight twin_plus i.b4.maternal_age i.parity i.b2.maternal_bmi smoker partner i.ie_type i.level dispo disp_ young old male if subject_grade==3 & level <= 2, cl(mor_id)

reg z_score i.byear i.treated i.b4.weight twin_plus i.b4.maternal_age i.parity i.b2.maternal_bmi smoker partner i.ie_type dispo disp_ young old male if subject_grade==3 & level > 2, cl(mor_id)
eststo e4
margins treated
reg z_score i.byear NC IUI i.b4.weight twin_plus i.b4.maternal_age i.parity i.b2.maternal_bmi smoker partner i.ie_type dispo disp_ young old male if subject_grade==3 & level > 2, cl(mor_id)

esttab e* using $data/tables/grade2_3edu.rtf, plain par b(3) ci(3) label replace mtitle("Low edu, grade 2" "High edu, grade 2" "Low edu, grade 3" "High edu, grade3")
```
